# Leading Cigarette Brands in the United States (2013-2023) by Brand Tier and Menthol Status

**DOI:** 10.64898/2025.12.08.25341575

**Authors:** Kathryn C. Edwards, Zhiqun Tang, Meagan O. Robichaud, Richard J. O’Connor, Lauren R. Pacek, Andrea C. Villanti, Cristine Delnevo

**Author notes:** **Corresponding Author:** Kathryn C. Edwards, PhD 1600 Research Blvd. Rockville, MD 20850.

## Abstract

**Importance:** While sales and use of cigarettes have declined, it is important to understand how these trends differ across brands and product characteristics, and how the demographic makeup of cigarette smokers has shifted.

**Objective:** Examine trends in U.S. market share for leading cigarette brands by brand tier and menthol status and describe the sociodemographic profile of people who use top cigarette brands.

**Design:** Cohort study: The Population Assessment of Tobacco and Heath (PATH) Study collects data from a nationally representative sample of U.S. youth (age 12-17) and adults (age 18+) in 1-2-year intervals from 2013-2023 (Waves 1-7).

**Setting:** Respondents complete the questionnaire via in-person audio-computer-assisted interviews. Telephone interviews were available in 2020-2023 due to the COVID-19 pandemic.

**Participants:** This study utilized data from 19,722 individuals (youth: N=1,201; adults: N=18,521) who smoked cigarettes in the past 30-days (P30D)

**Exposures:** Time (survey wave).

**Main Outcome and Measures:** Respondents were asked the cigarette brand that they usually smoke or last smoked, the number of cigarettes smoked per day, and how many days they smoked in the P30D to estimate monthly intake and market share. Cigarette brands were coded into premium and non-premium brand tiers. Demographic characteristics, other tobacco use, alcohol use, marijuana use, and mental health status were also assessed.

**Results:** Premium brands (overall and menthol) experienced significant market declines. Non-premium brands saw a market share increase. Characteristics of those smoking cigarettes changed: reduced number of cigarettes smoked per day, increased marijuana use, decreased alcohol use, and decreased moderate-to-high-severity mental health symptoms.

**Conclusions and Relevance:** While brand loyalty remains strong for the top three cigarette brands, the expanding non-premium market continues to encroach on the premium brand market. Similar patterns were observed in the menthol sector. As the marketplace changes so do the profiles of the people who use them or market potentially reflects the people who are still smoking. Continued timely surveillance of cigarette brand preferences and profiles of the people who use them will inform tobacco control policies that minimize tobacco-related mortality.

**Key Points:** *Objective:* Examine market share trends (2013-2023) for cigarette brands by brand tier and menthol status; describe the sociodemographic profile of people who use top cigarette brands.

*Findings:* Premium brands experienced significant market declines overall and among the menthol category. Non-premium brands saw a market share increase. Characteristics of those smoking cigarettes changed: reduced cigarettes smoked per day, increased marijuana use, decreased alcohol use, and decreased moderate-to-high-severity mental health symptoms.

*Meaning:* While brand loyalty remains strong for top cigarette brands, the expanding non-premium market is encroaching on the premium brand market. These changes reflect shifts in sociodemographic profiles of people smoking in 2023.

## Introduction

While overall sales and use of cigarettes have declined over the past 25 years in the United States (U.S.),^1-4^ these trends have differed across brands and product characteristics, and the demographic makeup of cigarette smokers has shifted.^5-10^ Factors impacting sales include restrictions on cigarette advertising, introduced by the Master Settlement Agreement (1998) and the Tobacco Control Act (TCA, 2009), preventing cigarette companies from advertising through most traditional marketing channels (e.g., billboards, sponsorship).^11^ Nevertheless, cigarettes continue to be one of the most heavily promoted products in the U.S. In 2022, tobacco companies spent $8.01 billion on cigarette advertising,^3^ primarily at the point-of-sale. Other factors influencing declining cigarette use are effective public health campaigns, the passage of smoke-free laws, and increased taxation.^12^ Taxation, in particular, may increase the appeal of “non-premium” or “discount” brands.^13^ One study found a non-significant increase in the proportion of smokers using non-premium brands (versus premium brands) between 2002 and 2011, with a trend toward more smokers switching from a premium to non-premium brand over time.^5^

Additionally, monitoring menthol cigarettes use trends remains important since their use is associated with increased smoking initiation, increased nicotine dependence, and decreased likelihood of sustained smoking cessation, particularly among individuals identifying as Black ^14-21^. Sales data and population studies show that menthol cigarette use has declined more slowly than non-menthol cigarette use, contributing to an increasing market share for menthol cigarettes ^3,6,8,22^. One study of Tobacco Tax and Trade Bureau data found that 91% of the decline in cigarette sales between 2009 and 2018 was attributable to non-menthol cigarettes, with use of non-menthol cigarettes declining by 33.1% (compared to 8.2% for menthol cigarettes ^8^). Since 2020, two states, Washington D.C., and a growing number of local jurisdictions have banned menthol cigarettes; therefore, current studies assessing trends in the use of menthol cigarettes are warranted ^23^.

Several factors shape consumers’ cigarette brand preferences, including product availability, economic circumstances, cigarette prices, tobacco marketing, and brand loyalty ^24^. Therefore, monitoring trends in use of different cigarette brands is important for detecting shifts in consumer preferences and can provide insight into the impacts of tobacco regulations and tobacco industry marketing.

However, data on such trends since the COVID-19 pandemic have been limited.^25^ Additionally, many studies that have examined demographic and substance use profiles of people who use leading cigarette brands did not examine trends across several demographic and substance use patterns often associated with different tobacco use patterns, including sexual orientation, use of non-cigarette tobacco/nicotine products (e.g., e-cigarettes), mental health status, and use of other substances (e.g., alcohol, marijuana). Using data from Waves 1-7 (W1-W7) of the Population Assessment of Tobacco and Health (PATH) Study (2013/2014-2022/2023), this study examines trends in market share for leading cigarette brands (non-menthol and menthol combined) in the U.S., and, separately among menthol brands alone. It also evaluates how the market shares for premium versus non-premium brands have shifted between 2013-2023. Lastly, this study analyzes the demographic, mental health status, and substance use profile of people who use leading cigarette brands.

## Methods

### Participants

The PATH Study is an ongoing, nationally representative, longitudinal cohort study of adults and youth in the U.S. that collects information on tobacco-use patterns and associated health behaviors. The PATH Study employed a stratified address-based, area-probability sampling design at W1 (2013/14) that oversampled adult tobacco users, young adults (aged 18-24 years), and Black adults. Further details regarding the PATH Study design and methods are published elsewhere ^26-29^. Details on interview procedures, questionnaires, sampling, weighting, response rates, and accessing the data are described in the <u>PATH Study Public Use Files User</u> <u>Guide Updated for Wave 7</u>.^30^ The study was conducted by Westat and approved by the Westat Institutional Review Board. All participants ages 18 and older provided informed consent, and youth participants aged 12–17 provided assent with each youth’s parent/legal guardian providing consent.

The present study utilized data from the public-use data files (PUF; https://doi.org/10.3886/ICPSR36498.v22, accessed and downloaded on 12/24/2024). We analyzed W1 to W7 data from youth (12-17 years old) and adults aged 18+ years who participated in the PATH Study and smoked cigarettes during the past 30-days.

### Cigarette Brand Measures

The analyses were based on a total of 19,722 unique respondents who were aged 12 and older (1,201 youth; 18,521 adults) and smoked cigarettes in the past 30-days (i.e., current smoking). No distinctions were made regarding single or poly-tobacco use (i.e., respondents who currently use cigarettes may have also used other tobacco products). Respondents were asked the cigarette brand that they usually smoke or last smoked. Sample sizes varied by wave and are presented in Supplemental Table 1.

Among these respondents, for each year between 2013/14 through 2022/23, we examined the frequency distribution of their regular brand and restricted our focus to the top 10 cigarette brands each year, resulting in a shortlist of 13 brands across the study period. This analysis was repeated for the subgroup who reported their usual/last brand was “menthol or mint” flavored, hereafter referred to as menthol. This resulted in a shortlist of 15 brands for the menthol subgroup. Cigarette brands were further coded into premium or non-premium brands as described in Supplemental Table 2.

### Demographic and Substance Use Measures

Demographic characteristics, other tobacco use, alcohol use, marijuana use, and mental health status (as measured via the GAIN-SS^31^) are described in Supplemental Table 3.

### Weight Adjustment

The PATH Study provides various weights at each wave to adjust for the complex sample design (e.g., oversampling of demographic groups) and nonresponse. We chose the appropriate weights for cross-sectional or pseudo-cross-sectional estimation at each wave (Supplemental Table 4).

Our analysis adjusted the weighting variables (one full-sample weight and 100 replicate weights per wave) for Tables 1-2 and Figures 1-2 by multiplying each of them by our derived past 30-day cigarette use measure (see Supplemental Table 3). The adjusted weights reflect the number of cigarettes consumed by current smokers in the past 30-days, accounting for use patterns among individuals and producing a better picture of the market in terms of the actual number of cigarettes smoked.^6^

**Figure 1.**
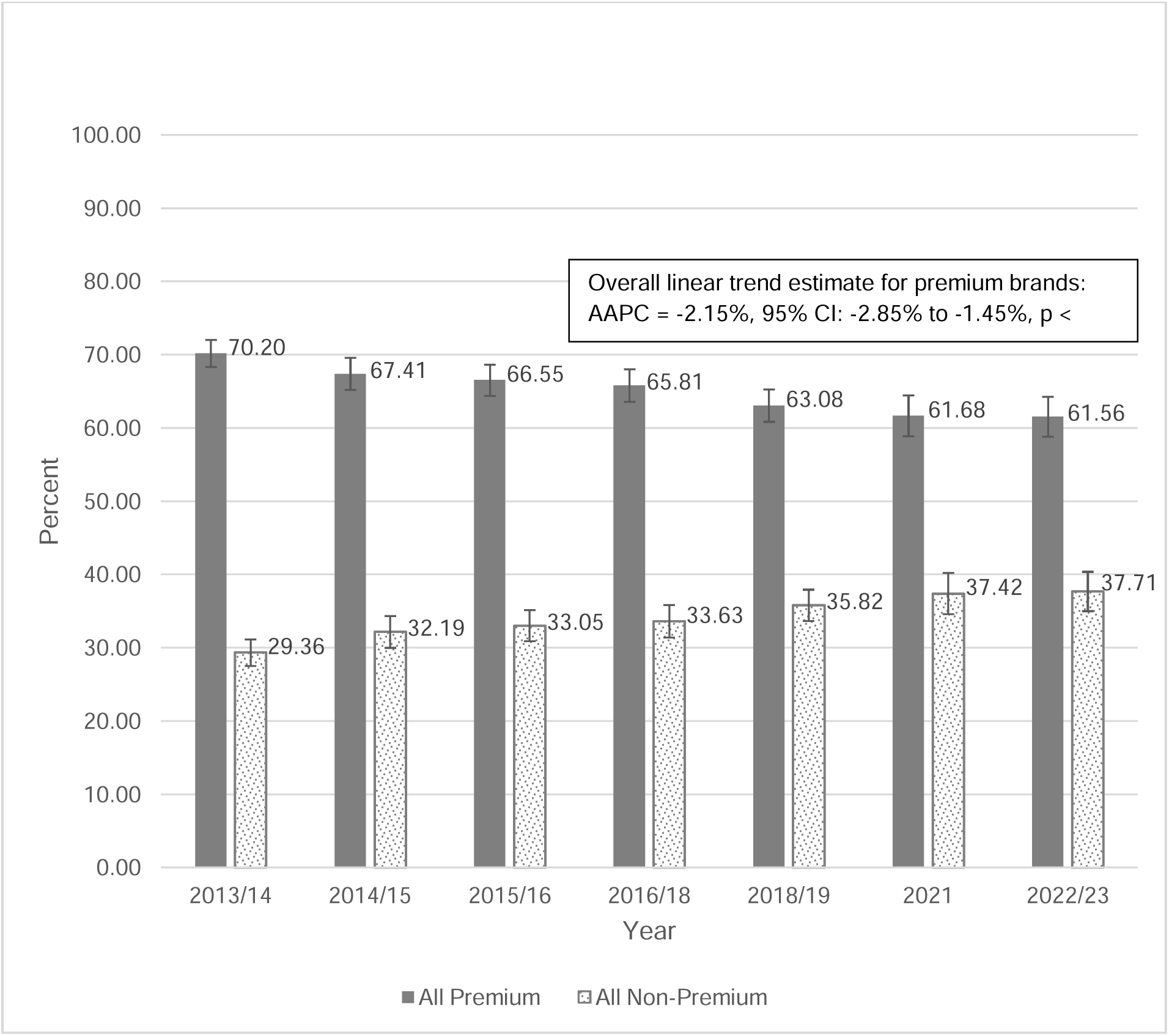
Market share of all brands: Premium vs. Non-Premium. Notes: Weighted percentages and weighted 95% Wilson confidence limits were presented. The total percentage of all brands by year was slightly smaller than 100% due to a handful of responses not being a cigarette brand (e.g., don’t know, refused, or a non-cigarette brand, which was <1% at each wave except for wave 5 in 2018/19, which was 1.1%). AAPC: Average Annual Percentage Change, estimated by WGEEs (with log link and non-premium brands as the reference).

**Figure 2.**
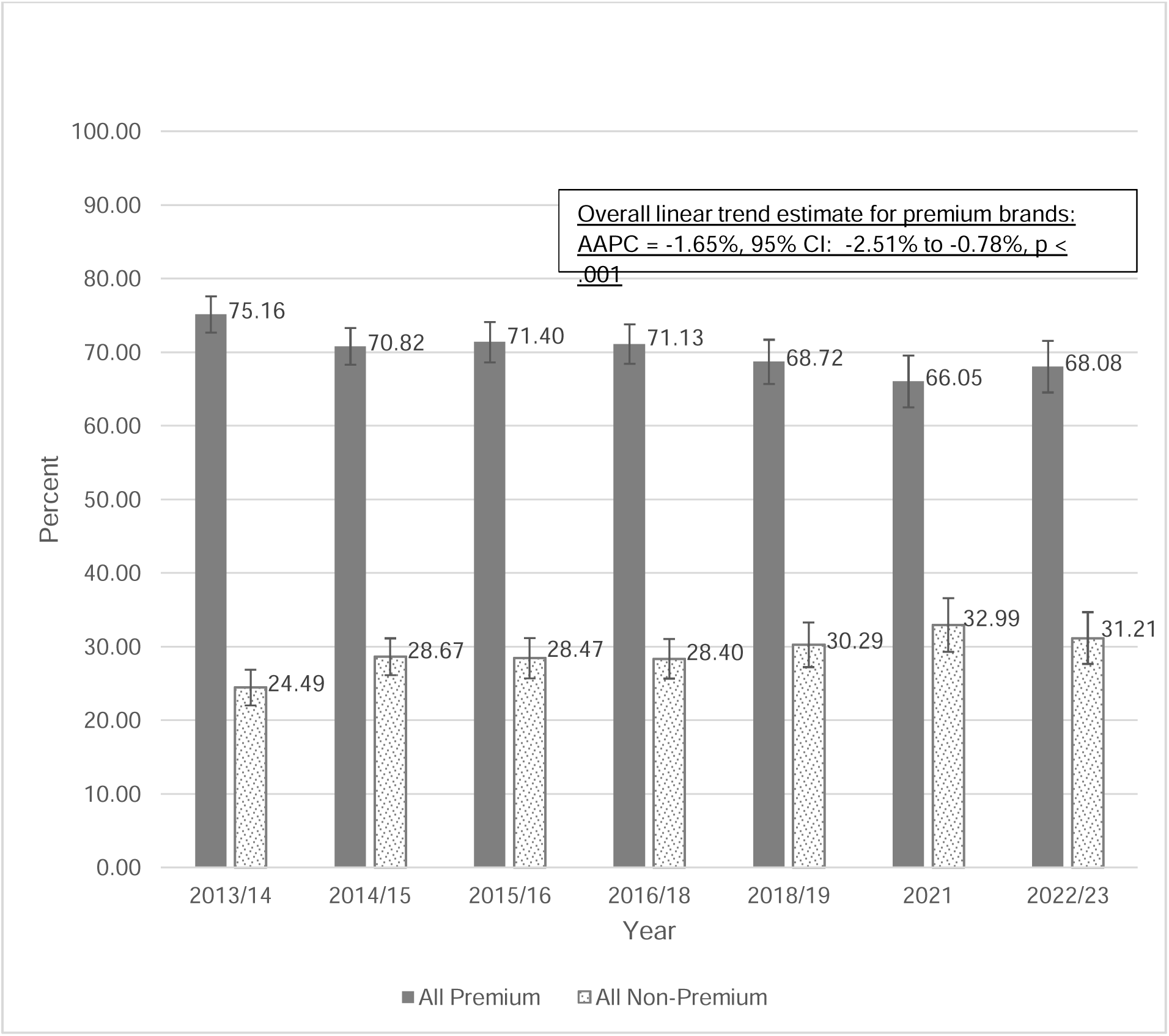
Market share of menthol styles: Premium vs. Non-Premium. Notes: Weighted percentages and weighted 95% Wilson confidence limits were presented. The total percentage of all brands by year was slightly smaller than 100% due to a handful of responses not being a cigarette brand (e.g., don’t know, refused, or a non-cigarette brand, which was <1% at each wave except for Wave 5 in 2018/19, which was 1.1%). AAPC: Average Annual Percentage Change, estimated by WGEEs (with log link and menthol style non-premium brands as the reference).

**Table 1.** Market share of all brands, regular or menthol, appearing in PATH’s top leading brand in the years 2013-2023 (Wave 1: 9/2013-12/2014 through Wave 7: 01/2022-04/2023)

| <u>Brand</u> | <u>2013/14</u> | <u>2014/15</u> | <u>2015/16</u> | <u>2016/18</u> | <u>2018/19</u> | <u>2021</u> | <u>2022/23</u> | <u>Change %</u><br><u>(2013-2023)</u> |
| --- | --- | --- | --- | --- | --- | --- | --- | --- |
| **Marlboro** | 39.26<br>(37.57,40.98) | 36.44<br>(34.61,38.31) | 35.74<br>(33.85,37.69) | 34.64<br>(32.81,36.52) | 32.22<br>(30.52,33.97) | 30.54<br>(28.51,32.64) | 30.39<br>(28.08,32.81) | -22.6 |
| **Newport** | 14.02<br>(12.64,15.53) | 13.54<br>(12.06,15.16) | 13.90<br>(12.53,15.40) | 14.11<br>(12.76,15.57) | 14.93<br>(13.45,16.54) | 14.95<br>(13.30,16.78) | 13.37<br>(11.64,15.31) | -4.7 |
| **Camel** | 7.98<br>(7.17, 8.89) | 9.43<br>(8.48,10.46) | 9.00<br>(8.07,10.02) | 8.98<br>(8.00,10.07) | 8.05<br>(7.08, 9.14) | 7.99<br>(6.79, 9.37) | 8.02<br>(6.73, 9.53) | .4 |
| Pall Mall | 8.49<br>(7.50, 9.60) | 9.15<br>(8.26,10.13) | 9.19<br>(8.22,10.26) | 8.70<br>(7.69, 9.83) | 8.79<br>(7.90, 9.77) | 7.57<br>(6.36, 8.99) | 7.62<br>(6.30, 9.19) | -10.3 |
| Maverick | 3.11<br>(2.47, 3.90) | 2.46<br>(1.90, 3.17) | 2.29<br>(1.73, 3.02) | 2.61<br>(2.00, 3.41) | 2.61<br>(1.95, 3.50) | 2.69<br>(1.81, 3.99) | 2.98<br>(2.19, 4.05) | -4.0 |
| L & M | 2.84<br>(2.35, 3.43) | 3.60<br>(2.94, 4.40) | 3.89<br>(3.26, 4.65) | 4.22<br>(3.52, 5.05) | 4.09<br>(3.17, 5.27) | 3.53<br>(2.65, 4.71) | 2.88<br>(2.23, 3.71) | 1.4 |
| Sonoma | 1.01<br>(0.55, 1.86) | 1.06<br>(0.55, 2.05) | 1.02<br>(0.61, 1.71) | 0.88<br>(0.46, 1.67) | 1.50<br>(0.92, 2.44) | 2.29<br>(1.27, 4.07) | 2.80<br>(1.69, 4.60) | 177.1 |
| 305's | 1.22<br>(0.80, 1.87) | 1.25<br>(0.81, 1.91) | 1.59<br>(1.03, 2.46) | 1.78<br>(1.15, 2.75) | 1.88<br>(1.22, 2.87) | 2.22<br>(1.40, 3.53) | 2.58<br>(1.54, 4.31) | 111.4 |
| **Kool** | 2.81<br>(2.19, 3.59) | 1.75<br>(1.29, 2.37) | 1.83<br>(1.37, 2.44) | 1.85<br>(1.40, 2.43) | 1.86<br>(1.31, 2.63) | 1.96<br>(1.48, 2.58) | 2.56<br>(1.87, 3.50) | -8.8 |
| **American Spirit** | 1.41<br>(1.11, 1.80) | 1.78<br>(1.48, 2.14) | 1.74<br>(1.38, 2.21) | 2.19<br>(1.84, 2.61) | 2.46<br>(2.01, 3.00) | 2.27<br>(1.77, 2.91) | 2.25<br>(1.84, 2.73) | 59.0 |
| **Winston** | 1.85<br>(1.53, 2.23) | 1.54<br>(1.26, 1.89) | 1.95<br>(1.49, 2.56) | 1.93<br>(1.51, 2.48) | 1.59<br>(1.22, 2.07) | 1.50<br>(1.10, 2.05) | 1.94<br>(1.45, 2.60) | 5.0 |
| Basic | 1.01<br>(0.77, 1.32) | 2.21<br>(1.79, 2.72) | 1.71<br>(1.28, 2.28) | 1.91<br>(1.54, 2.36) | 1.54<br>(1.05, 2.25) | 0.86<br>(0.45, 1.64) | 1.04<br>(0.66, 1.63) | 3.2 |
| Pyramid | 1.76<br>(1.26, 2.46) | 1.79<br>(1.41, 2.26) | 1.86<br>(1.43, 2.42) | 1.71<br>(1.34, 2.19) | 1.46<br>(1.09, 1.94) | 0.76<br>(0.49, 1.17) | 0.81<br>(0.48, 1.37) | -54.0 |
| <u>Summary among Top 10 Brands</u> |  |  |  |  |  |  |  |  |
| Top 10: Premium | 67.34<br>(65.46, 69.16) | 64.48<br>(62.35, 66.54) | 64.18<br>(62.05, 66.27) | 63.70<br>(61.47, 65.88) | 61.00<br>(58.83, 63.13) | 59.21<br>(56.38, 61.98) | 58.53<br>(55.71, 61.28) | -13.1 |
| Top 10: Non-Premium | 19.44<br>(18.01, 20.96) | 21.51<br>(20.07, 23.03) | 21.55<br>(20.27, 22.88) | 21.82<br>(20.39, 23.31) | 21.98<br>(20.37, 23.68) | 19.92<br>(18.09, 21.88) | 20.71<br>(18.55, 23.06) | 6.6 |
| All Top 10 | 86.78<br>(85.08, 88.31) | 85.99<br>(83.92, 87.83) | 85.73<br>(83.91, 87.38) | 85.52<br>(83.93, 86.97) | 82.98<br>(81.11, 84.71) | 79.12<br>(76.72, 81.35) | 79.24<br>(76.90, 81.40) | -8.7 |
| <u>Summary among All Brands</u> |  |  |  |  |  |  |  |  |
| All Premium | 70.20<br>(68.30, 72.02) | 67.41<br>(65.19, 69.55) | 66.55<br>(64.39, 68.65) | 65.81<br>(63.55, 68.00) | 63.08<br>(60.84, 65.26) | 61.68<br>(58.85, 64.43) | 61.56<br>(58.79, 64.25) | -12.3 |
| All Non-Premium | 29.36<br>(27.56, 31.22) | 32.19<br>(30.04, 34.41) | 33.05<br>(30.95, 35.21) | 33.63<br>(31.46, 35.88) | 35.82<br>(33.70, 37.99) | 37.42<br>(34.64, 40.29) | 37.71<br>(35.05, 40.44) | 28.4 |
| All Brands<sup>#</sup> | 99.55 | 99.60 | 99.60 | 99.44 | 98.90 | 99.10 | 99.27 | -0.3 |

Notes: Focused on the top 10 cigarette brands each year, resulting in a shortlist of 13 brands across the entire time period. Weighted percentages with Wilson confidence limits were presented. The top 10 brands at any wave were ordered by Wave 7, and **bolded are premium brands**. Analyses did not include respondents who only used roll-your-own cigarette tobacco.
<sup>#</sup>The total weighted percentage of all brands by year was slightly smaller than 100% due to small number of responses not being a cigarette brand (e.g., don't know, refused, or a non-cigarette brand).

**Table 2.** Market share of brands’ menthol/mint styles appearing in PATH’s Top 10 in the years 2013-2023 (Wave 1: 9/2013-12/2014 through Wave 7: 01/2022-04/2023)

| <u>Brand</u> | <u>2013/14</u> | <u>2014/15</u> | <u>2015/16</u> | <u>2016/18</u> | <u>2018/19</u> | <u>2021</u> | <u>2022/23</u> | <u>Change %</u><br><u>(2013-2023)</u> |
| --- | --- | --- | --- | --- | --- | --- | --- | --- |
| **Newport** | 35.09<br>(32.12,38.18) | 27.93<br>(25.24,30.79) | 29.09<br>(26.61,31.70) | 28.67<br>(26.11,31.39) | 29.22<br>(26.36,32.26) | 31.62<br>(28.46,34.97) | 28.62<br>(25.23,32.28) | -18.4 |
| **Marlboro** | 19.67<br>(17.98,21.46) | 22.96<br>(20.78,25.30) | 24.40<br>(22.12,26.84) | 23.04<br>(20.83,25.41) | 20.17<br>(18.26,22.22) | 17.55<br>(15.47,19.84) | 18.38<br>(15.83,21.24) | -6.5 |
| **Camel** | 8.49<br>(7.33, 9.82) | 11.02<br>(9.94,12.20) | 9.91<br>(8.75,11.21) | 10.85<br>(9.17,12.80) | 10.40<br>(8.97,12.03) | 8.52<br>(6.76,10.70) | 9.43<br>(7.20,12.25) | 11.0 |
| Pall Mall | 5.52<br>(4.38, 6.92) | 7.56<br>(6.24, 9.13) | 7.65<br>(6.24, 9.34) | 7.30<br>(5.96, 8.90) | 7.29<br>(5.77, 9.16) | 6.08<br>(4.36, 8.44) | 6.28<br>(4.20, 9.30) | 13.9 |
| **Kool** | 7.37<br>(5.70, 9.47) | 3.79<br>(2.61, 5.47) | 3.96<br>(2.95, 5.29) | 4.34<br>(3.34, 5.63) | 4.31<br>(3.09, 5.98) | 4.19<br>(3.04, 5.74) | 6.19<br>(4.52, 8.42) | -16.0 |
| Maverick | 5.46<br>(4.19, 7.07) | 4.12<br>(3.04, 5.57) | 2.78<br>(2.04, 3.77) | 3.64<br>(2.71, 4.88) | 3.85<br>(2.57, 5.75) | 3.27<br>(2.40, 4.44) | 3.58<br>(2.52, 5.07) | -34.4 |
| L & M | 2.06<br>(1.57, 2.71) | 3.52<br>(2.65, 4.66) | 3.47<br>(2.74, 4.38) | 4.18<br>(3.27, 5.33) | 4.32<br>(3.25, 5.71) | 3.93<br>(2.48, 6.16) | 3.25<br>(2.45, 4.29) | 57.6 |
| Seneca | 0.53<br>(0.20, 1.37) | 0.87<br>(0.33, 2.26) | 1.29<br>(0.59, 2.78) | 1.24<br>(0.57, 2.70) | 1.65<br>(0.85, 3.21) | 2.12<br>(1.24, 3.63) | 2.46<br>(1.20, 4.95) | 362.2 |
| Sonoma | 0.84<br>(0.45, 1.57) | 1.04<br>(0.50, 2.15) | 1.03<br>(0.53, 1.97) | 0.75<br>(0.32, 1.76) | 1.14<br>(0.59, 2.19) | 1.65<br>(0.80, 3.37) | 2.09<br>(1.17, 3.68) | 146.8 |
| 305's | 0.86<br>(0.56, 1.33) | 1.09<br>(0.67, 1.78) | 1.07<br>(0.66, 1.74) | 1.26<br>(0.73, 2.15) | 1.55<br>(0.96, 2.47) | 2.40<br>(1.34, 4.27) | 2.06<br>(1.13, 3.72) | 138.9 |
| Doral | 1.08<br>(0.69, 1.71) | 0.40<br>(0.21, 0.76) | 0.50<br>(0.26, 0.97) | 0.29<br>(0.11, 0.77) | 0.58<br>(0.29, 1.15) | 0.36<br>(0.16, 0.81) | 1.15<br>(0.53, 2.44) | 5.7 |
| **Salem** | 2.31<br>(1.68, 3.16) | 1.59<br>(1.12, 2.26) | 1.61<br>(1.13, 2.29) | 1.82<br>(1.31, 2.52) | 1.56<br>(0.95, 2.57) | 1.23<br>(0.76, 1.99) | 1.14<br>(0.60, 2.16) | -50.5 |
| Misty | 1.49<br>(1.07, 2.07) | 1.27<br>(0.84, 1.94) | 1.20<br>(0.76, 1.88) | 1.58<br>(1.06, 2.35) | 1.16<br>(0.71, 1.88) | 0.92<br>(0.52, 1.65) | 1.07<br>(0.55, 2.07) | -28.1 |
| USA Gold | 0.88<br>(0.47, 1.62) | 1.39<br>(0.82, 2.35) | 0.94<br>(0.52, 1.68) | 0.84<br>(0.44, 1.58) | 0.93<br>(0.48, 1.81) | 0.94<br>(0.50, 1.75) | 0.87<br>(0.43, 1.75) | -1.0 |
| Basic | 1.01<br>(0.66, 1.54) | 1.82<br>(1.27, 2.59) | 1.42<br>(0.90, 2.25) | 1.43<br>(1.05, 1.93) | 1.17<br>(0.63, 2.18) | 0.46<br>(0.18, 1.17) | 0.37<br>(0.19, 0.73) | -63.6 |
| <u>Summary among Top 10 Brands</u> |  |  |  |  |  |  |  |  |
| Top 10: Premium | 72.92<br>(70.51, 75.20) | 67.30<br>(64.82, 69.70) | 68.98<br>(66.21, 71.62) | 68.72<br>(66.01, 71.30) | 65.67<br>(62.49, 68.70) | 63.10<br>(59.38, 66.68) | 63.76<br>(59.89, 67.46) | -12.6 |
| Top 10: Non-Premium | 19.74<br>(17.60, 22.07) | 23.09<br>(20.95, 25.37) | 21.32<br>(19.19, 23.63) | 22.52<br>(20.22, 25.00) | 23.65<br>(21.17, 26.32) | 22.14<br>(19.47, 25.07) | 23.17<br>(20.21, 26.42) | 17.4 |
| All Top 10 | 92.65<br>(91.24, 93.85) | 90.39<br>(88.15, 92.24) | 90.31<br>(88.13, 92.12) | 91.24<br>(89.84, 92.46) | 89.31<br>(87.38, 90.98) | 85.25<br>(81.80, 88.14) | 86.93<br>(84.42, 89.10) | -6.2 |
| <u>Summary among All Brands</u> |  |  |  |  |  |  |  |  |
| All Premium | 75.16<br>(72.64, 77.52) | 70.82<br>(68.26, 73.26) | 71.40<br>(68.57, 74.07) | 71.13<br>(68.40, 73.72) | 68.72<br>(65.63, 71.65) | 66.05<br>(62.45, 69.48) | 68.08<br>(64.46, 71.48) | -9.4 |
| All Non-Premium | 24.49<br>(22.15, 26.99) | 28.67<br>(26.22, 31.24) | 28.47<br>(25.81, 31.29) | 28.40<br>(25.78, 31.18) | 30.29<br>(27.34, 33.41) | 32.99<br>(29.45, 36.75) | 31.21<br>(27.80, 34.84) | 27.4 |
| All Brands<sup>#</sup> | 99.65 | 99.49 | 99.87 | 99.53 | 99.01 | 99.05 | 99.28 | -0.4 |

Notes: Focused the top 10 cigarette brands for those who use menthol/mint, resulting in a shortlist of 15 brands across the entire time period. Weighted percentages with Wilson confidence limits were presented. The top 10 brands at any wave were ordered by Wave 7, and **bolded are premium brands**. Analyses did not include respondents who only used roll-your-own cigarette tobacco.
<sup>#</sup>The total weighted percentage of all brands by year was slightly smaller than 100% due to a handful of responses not being a cigarette brand (e.g., don't know, refused, or a non-cigarette brand).

### Statistical Analysis

Applying the adjusted weight variables, we ran frequency distributions of the “usually/last smoked brand” variable for each year individually, overall, and for menthol. We also examined changes in market share between 2013/14 and 2022/23 by computing the percentage change in market share for each brand across nearly 10 years and tested differences in market shares (e.g., all premium versus all non-premium brands) in 2023 versus 2013, using weighted Generalized Estimating Equations (WGEEs) logistic regression analyses. A significant regression coefficient for the WAVE variable represents significant change from W1 to W7 (i.e., between 2013/14 and 2022/23), and an odds ratio (OR) >1 represents a positive change (i.e., increase from 2013/14 to 2022/23) and an OR <1 represents a decrease. We further used WGEEs (with log link) to obtain Average Annual Percent Change (AAPC), a summary measure of an observed trend (equivalent to a linear slope parameter) across the entire period.

We also examined demographic characteristics, tobacco use characteristics, other substance use (i.e., alcohol and marijuana) and mental health status among participants who used the top 10 cigarette brands at 2022/23, as well as overall and among premium versus non-premium brands (see Supplemental Table 2 for details about measures and variable definitions). For these analyses we used the original PATH Study weights described in Supplemental Table 4, without further adjusting by the cigarette use measure. To examine bivariate associations between a binary or continuous variables and the WAVE (i.e., survey year) variable (overall, all premium brands, and all non-premium brands) we also utilized WGEEs logistic regression analyses (Edwards, Kasza, Tang et al., 2020). GEEs (^32,33^) allow for inclusion of repeated measures in analysis while statistically controlling for dependence among observations contributed by the same individuals and can assess change across time. For categorical variables with 3 or more categories or levels, we used Rao-Scott Chi-Square tests. Rao-Scott Chi-Square tests were also used for a binary variable that WGEEs could not run through all the BRR weights. All analyses were conducted using SAS V.9.4 software (SAS Institute, Cary, North Carolina).

## Results

### Premium versus non-premium brands’ market share for 2013-2023

Figure 1 shows the breakdown of all premium versus all non-premium brands from 2013-2023. Overall, the market share for premium brands decreased, while that of non-premium brands increased over this ten-year period. The market share differences between all premium versus all non-premium brands in 2023 versus 2013 was significant (OR=0.72, 95%CI: 0.64 to 0.80, p<0.0001). The overall declining trend of all premium from 2013-2023, compared to all non-premium brands, was significant (AAPC = -2.15%, 95% CI: - 2.85% to -1.45%, p< 0.0001).

### Leading cigarette brands’ market share for 2013-2023

Table 1 shows market shares for leading cigarette brands for 2013-2023, as well as market shares for premium brands combined (leading, non-leading, and total) and non-premium brands combined (leading, non-leading, and total). While Marlboro remained the market leader (30.4% in 2023), their market share declined by approximately 23% over the ten-year period. Newport remained in the top two (13.4% in 2023) during this period with a more modest 4.7% decline. While Pall Mall (non-premium) entered the top three in 2013 (8.5% market share), edging out Camel (8.0%), they experienced a greater decline in market share over the study period (-10.3%) compared to Camel, which remained relatively stable (+0.4%). Non-premium brand Pyramid, saw their share of the market decline (-54.0%), while non-premium brands Sonoma and 305’s experienced large market share increases (+177.1% and +111.4%, respectively). Premium brand, American Spirt, also saw moderate growth (+59.0%). While the top ten brands from 2023 (Marlboro through American Spirit in Table 1) represented 82.2% of the market share in 2013, by 2023 they comprised 75.5% of the market.

This is reflected in the greater percentage reporting other brands (12.8% in 2013 versus 20.0% is 2023).

### Premium versus non-premium brands’ market share of menthol styles for 2013-2023

Figure 2 shows the breakdown of all menthol premium versus all non-premium brands from 2013-2023. During this period, premium brands exhibited decreasing market share, while non-premium brands exhibited an increase in market share, though there was a slight, non-significant, rebound from 2021-2023 for premium brands in the market. These trends resulted in a market share difference between premium versus non-premium menthol brands that was significantly smaller in 2023 compared to 2013 (OR=0.77, 95%CI: 0.65 to 0.92, p=0.0032). The overall declining trend of all menthol premium from 2013-2023, compared to all menthol style non-premium brands, was significant (AAPC = -1.65%, 95% CI: -2.51% to -0.78%, p<0.001).

### Leading cigarette brands’ market share of menthol styles for 2013-2023

Table 2 shows the leading cigarette brands’ market share of menthol styles for 2013-2023. Top brands in the menthol category continue to be premium brands Newport and Marlboro, though Newport’s market share declined by about 18% over the ten-year period (from 35.1% to 28.6%). Marlboro had a more modest 6.5% decline (from 19.7% to 18.4%), retaining the second largest market share in 2023. Among premium brands, Camel—with a market share of 8.5% in 2013 and 9.4% in 2023—remained in third place over the study period and was the only leading premium brand to that gained (+11.0%) market share. Market shares for two other premium brands, Kool and Salem, declined (-16.0% and -50.5%, respectively).

Conversely, a greater number of non-premium menthol brands experienced market share increases. Among them Pall Mall, which, like Camel, remained relatively stable in its market share ranking (fourth place over the study period) and gained market share (13.9%). Seneca, Sonoma, and 305’s—three small non-premium brands—experienced large market share increases (+362.2%, +146.8%, and +138.9%, respectively) over the study period. The brand with the largest loss was Basic (-63.6%), a non-premium brand.

### Demographic, tobacco, and substance use profile of people who smoke cigarettes during 2013-2023

Table 3 presents demographic, substance use profile (i.e., other tobacco/nicotine products, alcohol, and marijuana), and mental health status of people who smoke cigarettes, overall and by brand tier. Supplemental table 5 provides these results for the leading brands from 2023, and are discussed in the Supplemental material.

**Table 3.**
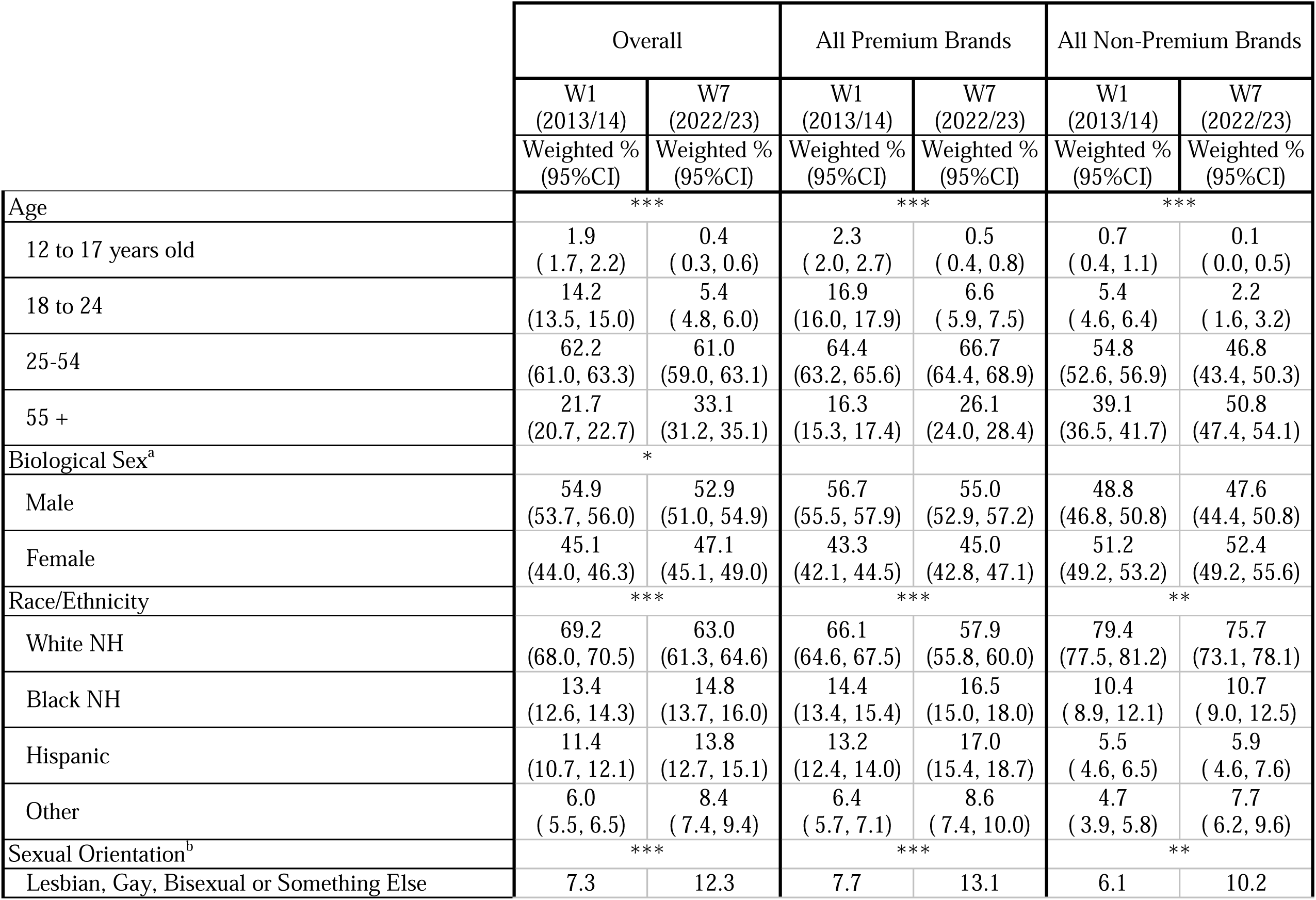
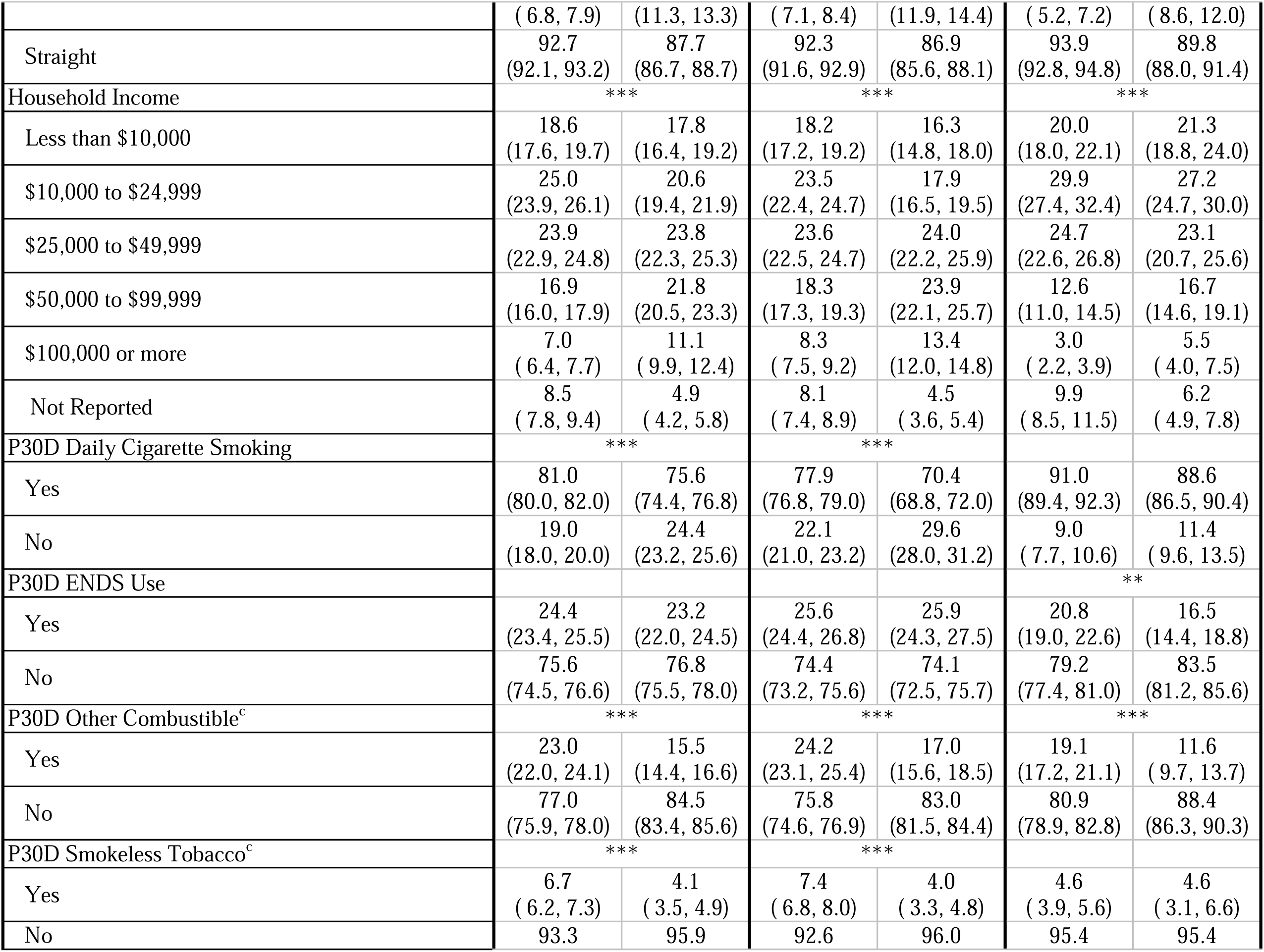
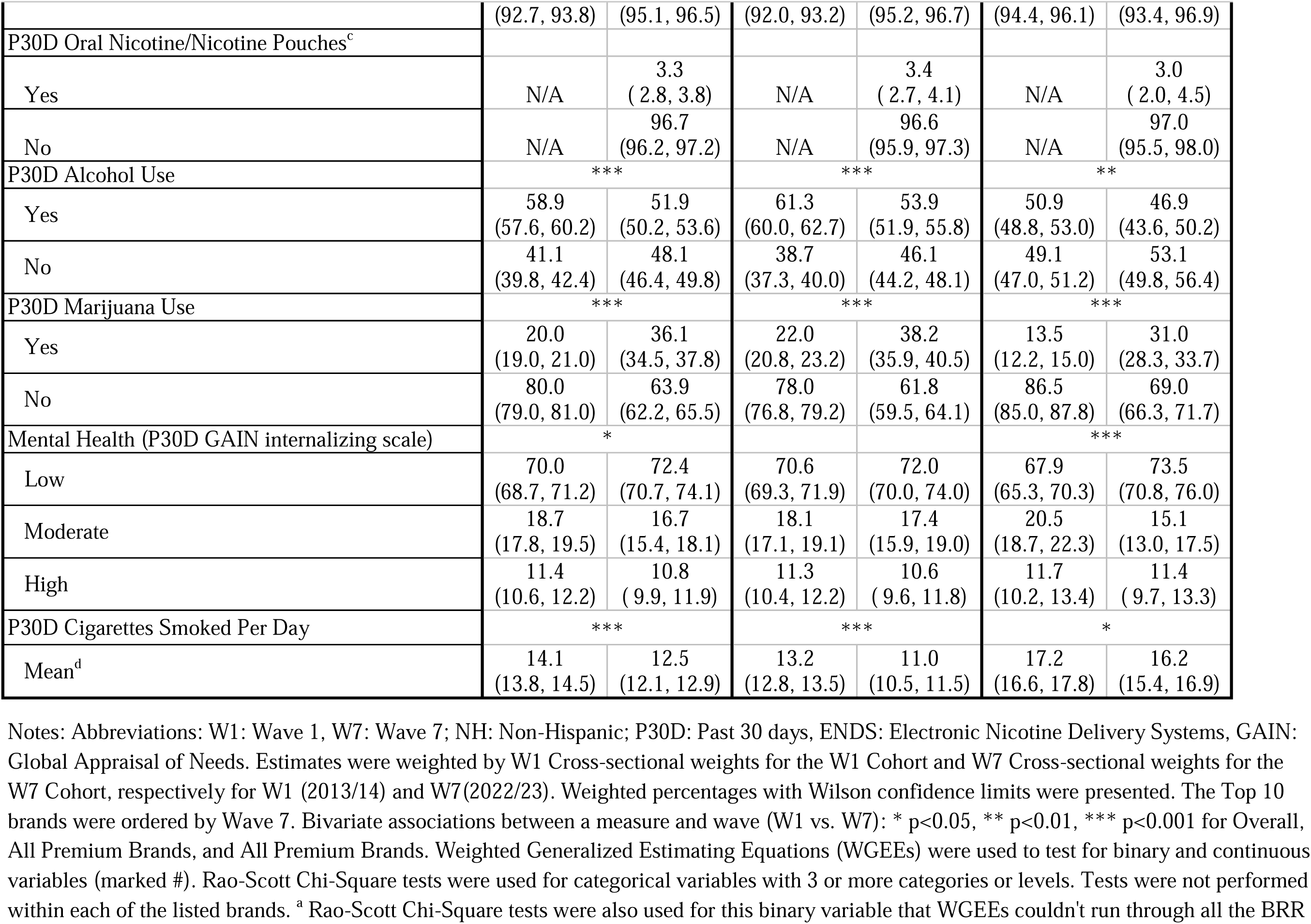
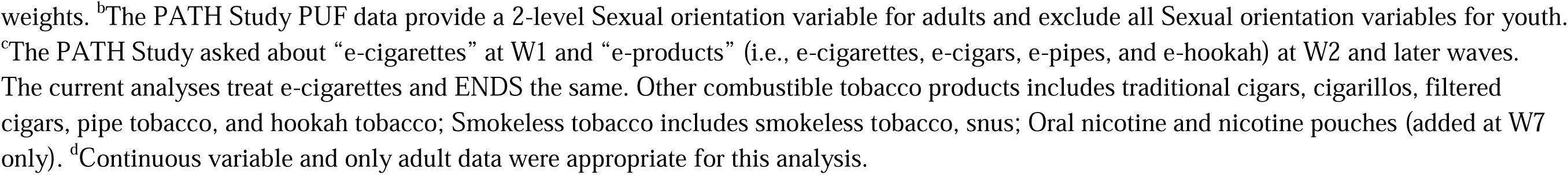
Demographic and substance use profile of those who use premium brands, non-premium brands and overall, 2013/14 and 2022/23.

#### Overall and by premium status (Table 3)

Overall, a higher proportion of individuals in 2023 (compared to 2013) were 55 or older, identified as a sexual minority (lesbian, gay, bisexual, or another sexual orientation), and had a household income of $50,000 or more, while a lower proportion were youth (12-17) or young adults (18-24) and individuals identifying as Non-Hispanic White (p<0.05). A lower proportion of individuals reported smoking daily, using other combustible products, and using smokeless tobacco in the past 30 days, and mean number of cigarettes smoked per day in the past 30 days decreased. Additionally, there were significant increases in past 30-day marijuana use but decreases in past 30-day alcohol use. Those experiencing moderate or high-severity mental health symptoms also decreased. These same patterns were present for people who used premium brands, except for sex and mental health symptoms, which were not significantly different between 2013 versus 2023. In contrast, among individuals using non-premium brands, there was a significant decrease in past-30-day ENDS use and moderate-severity mental health symptoms but no significant changes with respect to sex, use of smokeless tobacco, and daily smoking. Compared to the overall sample and people who smoke premium brands, people smoking non-premium brands smoked more cigarettes per day, and a higher proportion were 55 or older, female, identified as Non-Hispanic White, had a household income of less than $25,000, and reported daily smoking.

## Discussion

While overall cigarette use has decreased over time, our findings show that premium cigarette brands such as Marlboro, Newport, and Camel remain popular, though Marlboro and Newport saw their market share decline between 2013-2023, including those reported as a menthol brand. This may reflect a shift from use of premium brands to non-premium brands or increased quitting/reduced initiation with premium brands. Overall, premium brand market share declined 12% while non-premium brand market share increased 28% during the study period. In June 2025, an article from *Convenience Store News* stated that deep discount cigarettes were the only cigarette price tier segment that has not experienced a decline in sales volume, in contrast to the “super-premium,” premium, and discount segments^34^. Future research could explore the differences in the non-premium sector by deep discount versus discount.

This pattern of results by price tier segment was also apparent when limiting the analyses to those who report use of menthol brands, with a 9% decline in market share for premium menthol brands and a 27% increase in non-premium menthol brands. Menthol cigarettes continue to exacerbate tobacco-related health disparities^16,35^. Due to decades of targeted marketing by the tobacco industry, certain populations—including non-Hispanic Black and Native Hawaiian and Pacific Islander individuals, youth, and LGBTQ+ individuals—use menthol cigarettes at higher rates^16^. While more people who identified as Black used traditional menthol brands like Newport and Kool (consistent with Miller Lo et al.^6^), we did not see higher proportions of youth or sexual minorities among these menthol brand profiles as reported in prior studies^36,37^. However, we did see a higher proportion of other combusted tobacco use among those using Newport and Kool, likely cigar use^22,37-39^ potentially increasing the negative health outcomes due to dual combusted tobacco product use.^40^

In addition to demographic differences in menthol cigarette use, young adults, low-income individuals, and people with serious mental illnesses, smoke at higher rates compared to the general population^16,35,41^. While cigarette use among young adults declined overall during the study period, the largest proportion of young adult use was with brands American Spirit and Camel. American Spirit advertising has been shown to reduce harm perceptions of cigarettes which may result in increased use of this brand among vulnerable populations like youth and young adults.^42^ Examining trends in cigarette brand preferences across different demographic groups over time can be helpful for understanding how regulations and marketing practices may be impacting populations differently. Overall, people who smoke cigarettes in 2023 are older, more racially diverse, more likely to identify as lesbian/gay/bisexual, fewer have moderate to severe mental health issues, and a greater current marijuana use than those who smoked cigarettes in 2013.

A strength of this paper was its longitudinal data collection for a ten-year period that included not only key tobacco use characteristics, but other demographic (e.g., sexual orientation), mental health, and substance use measures absent in previous research. We summarize the data by premium versus non-premium status as well as report out the leading brands throughout the study period. Our research is limited to those who reported a usual/last-used brand, and while we account for cigarette use patterns (number of cigarettes smoked per month) in our market share estimates, we do not capture brand switching or estimate trajectories of use patterns. Future research could explore multivariable models to build upon our descriptive work and assess associations between demographic/substance use/mental health variables and cigarette use patterns.

In conclusion, while brand loyalty remains strong for the top three cigarette brands, the expanding non-premium market continues to encroach on the premium brand market. Premium brands that were present in 2014-2019^6^ were no longer seen in 2013-2023. This pattern is consistent in the menthol sector as well. As the marketplace changes so do the profiles of the people who use them or the market begins to reflect the people who are still smoking. Continued timely surveillance of cigarette brand preferences and profiles of the people who use them will provide regulators with data to make informed decisions for the protection of public health.

## Data Availability

This study used ONLY openly available data from the PATH Study Public Use Files available at: https://www.icpsr.umich.edu/web/NAHDAP/studies/36498

## References

1. Ali FRM, Seaman EL, Schillo B, Vallone D. Trends in Annual Sales and Pack Price of Cigarettes in the US, 2015-2021. JAMA Network Open. 2022;5(6):e2215407-e2215407. doi:10.1001/jamanetworkopen.2022.15407

2. Ganz O, LaVake M, Hrywna M, King Jensen JL, Delnevo CD. National Trends in Sales and Price for Commercial Tobacco and Nicotine Products, 2018-2022. JAMA Network Open. 2024;7(3):e241384-e241384. doi:10.1001/jamanetworkopen.2024.1384

3. US Federal Trade Commission. Cigarette Report for 2022 (2023). https://www.ftc.gov/system/files/ftc_gov/pdf/2022-Cigarette-Report.pdf

4. Cornelius ME LC, Jamal A, et al. Tobacco Product Use Among Adults – United States, 2021. Morbidity and Mortality Weekly Reports. 2023;72:475-483. doi: 10.15585/mmwr.mm7218a1

5. Cornelius ME, Driezen P, Fong GT, et al. Trends in the use of premium and discount cigarette brands: findings from the ITC US Surveys (2002-2011). Tob Control. Mar 2014;23 Suppl 1(0 1):i48-53. doi:10.1136/tobaccocontrol-2013-051045

6. Miller Lo EJ, Young WJ, Ganz O, Talbot EM, O’Connor RJ, Delnevo CD. Trends in Overall and Menthol Market Shares of Leading Cigarette Brands in the USA: 2014–2019. International Journal of Environmental Research and Public Health. 2022;19(4):2270.

7. Sharma A, Fix BV, Delnevo C, Cummings KM, O’Connor RJ. Trends in market share of leading cigarette brands in the USA: national survey on drug use and health 2002–2013. BMJ Open. 2016;6(1):e008813. doi:10.1136/bmjopen-2015-008813

8. Delnevo CD, Giovenco DP, Villanti AC. Assessment of Menthol and Nonmenthol Cigarette Consumption in the US, 2000 to 2018. JAMA Network Open. 2020;3(8):e2013601-e2013601. doi:10.1001/jamanetworkopen.2020.13601

9. Meza R, Cao P, Jeon J, Warner KE, Levy DT. Trends in US Adult Smoking Prevalence, 2011 to 2022. JAMA Health Forum. Dec 1 2023;4(12):e234213. doi:10.1001/jamahealthforum.2023.4213

10. Campaign for Tobacco Free Kids. TOBACCO USE AMONG AFRICAN AMERICANS. Accessed July 8, 2025, https://www.tobaccofreekids.org/fact-sheets/health-equity-and-special-populations/toll-of-tobacco-on-specific-populations-other-populations

11. Truth Initiative. What do tobacco advertising restrictions look like today? Accessed July 8, 2025, https://truthinitiative.org/research-resources/tobacco-industry-marketing/what-do-tobacco-advertising-restrictions-look-today

12. Pierce JP, White VM, Emery SL. What public health strategies are needed to reduce smoking initiation? Tob Control. Mar 2012;21(2):258–64. doi:10.1136/tobaccocontrol-2011-050359

13. Chaloupka FJ, Cummings KM, Morley CP, Horan JK. Tax, price and cigarette smoking: evidence from the tobacco documents and implications for tobacco company marketing strategies. Tob Control. Mar 2002;11 Suppl 1(Suppl 1):I62-72. doi:10.1136/tc.11.suppl_1.i62

14. Cook S, Hirschtick JL, Patel A, et al. A longitudinal study of menthol cigarette use and smoking cessation among adult smokers in the US: Assessing the roles of racial disparities and E-cigarette use. Prev Med. Jan 2022;154:106882. doi:10.1016/j.ypmed.2021.106882

15. Campaign for Tobacco Free Kids. IMPACT OF MENTHOL CIGARETTES ON YOUTH SMOKING INITIATION AND HEALTH DISPARITIES. Accessed July 8, 2025, https://assets.tobaccofreekids.org/factsheets/0390.pdf

16. Centers for Disease Control and Prevention. Menthol Smoking and Related Health Disparities. Accessed July 8, 2025, https://www.cdc.gov/tobacco/menthol-tobacco/health-disparities.html

17. Leas EC, Benmarhnia T, Strong DR, Pierce JP. Effects of menthol use and transitions in use on short-term and long-term cessation from cigarettes among US smokers. Tob Control. Apr 2023;32(e1):e31–e36. doi:10.1136/tobaccocontrol-2021-056596

18. Villanti AC, Collins LK, Niaura RS, Gagosian SY, Abrams DB. Menthol cigarettes and the public health standard: a systematic review. BMC Public Health. Dec 29 2017;17(1):983. doi:10.1186/s12889-017-4987-z

19. Villanti AC, Johnson AL, Glasser AM, et al. Association of Flavored Tobacco Use With Tobacco Initiation and Subsequent Use Among US Youth and Adults, 2013-2015. JAMA Netw Open. Oct 2 2019;2(10):e1913804. doi:10.1001/jamanetworkopen.2019.13804

20. Villanti AC, Johnson AL, Halenar MJ, et al. Menthol and Mint Cigarettes and Cigars: Initiation and Progression in Youth, Young Adults and Adults in Waves 1–4 of the PATH Study, 2013-2017. Nicotine Tob Res. Aug 4 2021;23(8):1318-1326. doi:10.1093/ntr/ntaa224

21. Villanti AC, Johnson AL, Ambrose BK, et al. Flavored Tobacco Product Use in Youth and Adults: Findings From the First Wave of the PATH Study (2013-2014). Am J Prev Med. Aug 2017;53(2):139–151. doi:10.1016/j.amepre.2017.01.026

22. Delnevo CD, Giovenco DP, Villanti AC. Trends in menthol and non-menthol cigarette consumption in the US: 2009-2024. Tob Control. Jun 18 2025;doi:10.1136/tc-2025-059408

23. Campaign for Tobacco Free Kids. STATES & LOCALITIES THAT HAVE RESTRICTED THE SALE OF FLAVORED TOBACCO PRODUCTS. Accessed July 8, 2025, https://assets.tobaccofreekids.org/factsheets/0398.pdf

24. Sharma A, Fix BV, Delnevo CD, Cummings KM, O’Connor RJ. Situational and Demographic Factors in the Sudden Growth of Pall Mall, 2002-2014. Nicotine Tob Res. May 3 2018;20(6):775-778. doi:10.1093/ntr/ntx140

25. Asare S, Majmundar A, Islami F, et al. Changes in Cigarette Sales in the United States During the COVID-19 Pandemic. Annals of Internal Medicine. 2022/01/18 2021;175(1):141-143. doi:10.7326/M21-3350

26. Hyland A, Ambrose BK, Conway KP, et al. Design and methods of the Population Assessment of Tobacco and Health (PATH) Study. Tob Control. Jul 2017;26(4):371–378. doi:10.1136/tobaccocontrol-2016-052934

27. Tourangeau R, Yan T, Sun H, Hyland A, Stanton CA. Population Assessment of Tobacco and Health (PATH) reliability and validity study: selected reliability and validity estimates. Tob Control. Nov 2019;28(6):663–668. doi:10.1136/tobaccocontrol-2018-054561

28. Piesse A, Opsomer J, Dohrmann S, et al. Longitudinal Uses of the Population Assessment of Tobacco and Health Study. Tob Regul Sci. Jan 2021;7(1):3–16. doi:10.18001/trs.7.1.1

29. Opsomer JD, Dohrmann S, DiGaetano R, et al. Update to the design and methods of the PATH Study, Wave 4 (2016-2017). Tob Control. Oct 19 2024;33(6):733-738. doi:10.1136/tc-2022-057851

30. United States Department of Health and Human Services. National Institutes of Health. National Institute on Drug Abuse, and United States Department of Health and Human Services. Food and Drug Administration. Center for Tobacco Products. Data from: Population Assessment of Tobacco and Health (PATH) Study [United States] Public-Use Files. 2025. doi:10.3886/ICPSR36498.v23

31. Dennis ML, Chan YF, Funk RR. Development and validation of the GAIN Short Screener (GSS) for internalizing, externalizing and substance use disorders and crime/violence problems among adolescents and adults. Am J Addict. 2006;15 Suppl 1(Suppl 1):80-91. doi:10.1080/10550490601006055

32. Liang KY, Zeger SL. Longitudinal data analysis using generalized linear models. Biometrika. 1986;73(1):13–22. doi:10.1093/biomet/73.1.13

33. Hubbard AE, Ahern J, Fleischer NL, et al. To GEE or not to GEE: comparing population average and mixed models for estimating the associations between neighborhood risk factors and health. Epidemiology. Jul 2010;21(4):467–74. doi:10.1097/EDE.0b013e3181caeb90

34. Covino RM. Cigarette Category Weathers Highs & Lows. Convenience Store News. Accessed July 8, 2025, https://csnews.com/cigarette-category-weathers-highs-lows

35. Eliminating Tobacco-Related Disease and Death: Addressing Disparities—A Report of the Surgeon General (2024).

36. Ganz O, Delnevo CD. Cigarette Smoking and the Role of Menthol in Tobacco Use Inequalities for Sexual Minorities. Nicotine Tob Res. Oct 7 2021;23(11):1942–1946. doi:10.1093/ntr/ntab101

37. Villanti AC, Mowery PD, Delnevo CD, Niaura RS, Abrams DB, Giovino GA. Changes in the prevalence and correlates of menthol cigarette use in the USA, 2004-2014. Tob Control. Nov 2016;25(Suppl 2):ii14-ii20. doi:10.1136/tobaccocontrol-2016-053329

38. Sterling K, Fryer C, Pagano I, Jones D, Fagan P. Association between menthol-flavoured cigarette smoking and flavoured little cigar and cigarillo use among African-American, Hispanic, and white young and middle-aged adult smokers. Tobacco Control. 2016;25(Suppl 2):ii21-ii31. doi:10.1136/tobaccocontrol-2016-053203

39. Rath JM, Villanti AC, Williams VF, Richardson A, Pearson JL, Vallone DM. Correlates of current menthol cigarette and flavored other tobacco product use among U.S. young adults. Addict Behav. Nov 2016;62:35–41. doi:10.1016/j.addbeh.2016.05.021

40. Chang CM, Corey CG, Rostron BL, Apelberg BJ. Systematic review of cigar smoking and all cause and smoking related mortality. BMC Public Health. 2015/04/24 2015;15(1):390. doi:10.1186/s12889-015-1617-5

41. Watkins SL, Pieper F, Chaffee BW, Yerger VB, Ling PM, Max W. Flavored Tobacco Product Use Among Young Adults by Race and Ethnicity: Evidence From the Population Assessment of Tobacco and Health Study. J Adolesc Health. Aug 2022;71(2):226–232. doi:10.1016/j.jadohealth.2022.02.013

42. Gratale SK, Maloney EK, Sangalang A, Cappella JN. Influence of Natural American Spirit advertising on current and former smokers’ perceptions and intentions. Tobacco Control. 2018;27(5):498–504. doi:10.1136/tobaccocontrol-2017-053881

