## Supplemental Material and Tables for "Leading Cigarette Brands in the United States (2013-2023) by Brand Tier and Menthol Status"

Supplemental Material for “Leading Cigarette Brands in the United States (2013-2023) by Brand Tier and Menthol Status”

**Supplemental Methods:**

*Coding Premium and Non-premium Cigarette Brands (for Supplemental Table 2)*

Initial premium brands were selected from Cornelius et al.^5^ Expert reviews from authors (MR, RO, CD) provided feedback for brands not included in that paper, as well as re-classifying L&M and Eve into the non-premium category based on brand descriptions from parent company reports.^31,32^

**Supplemental Results:**

*Demographics, Tobacco/Substance Use, and Mental Health Profiles by Specific Brand (Supplemental Table 5):*

The majority of people that smoked each brand identified as Non-Hispanic White in both 2013 and 2023, except for Newport and Kool (predominately menthol brands), which had higher proportions of individuals identifying as non-Hispanic Black. The top three brands (Marlboro, Newport, and Camel) had higher proportions of individuals identifying Hispanic, likely contributing to the higher proportion of Hispanic individuals among smokers using premium cigarettes.

With the exception of predominately menthol brands (Newport and Kool), the leading premium brands (i.e., Marlboro, Camel, and American Spirit) had greater proportions of individuals with an annual household income of $50,000 or more. Notably, the proportion of individuals with household incomes of $50,000 or more was higher for each of these brands in 2023 compared to 2013, while the proportion of individuals with household incomes of less than $25,000 decreased.

Daily smoking was more common among those who reported smoking non-premium brands (e.g., Pall Mall, Maverick, L&M). However, except for two non-premium brands (Sonoma and 305's), the prevalence of daily smoking and number of cigarettes smoked per day decreased from 2013-2023 for all leading brands

The proportion of individuals using other combustible tobacco products was highest for predominately menthol brands (Newport and Kool). In contrast to other leading brands, prevalence of other combustible tobacco use was higher than e-cigarette use among people using Newport and Kool. While the prevalence of other combustible tobacco use declined for most brands between 2013-2023, prevalence of other combustible tobacco use remained relatively stable for Kool and L&M. Prevalence of past 30-day ENDS varied between approximately a third and a fifth among premium brands, with Camel and American Spirit having the largest proportion of ENDS use in 2023, and Kool having the lowest. ENDS use also varied among non-premium brands, with Pall Mall having the lowest percentage of individuals reporting ENDS use in 2023 and having the largest decrease in ENDS use from 2013-2023 among leading brands. In contrast, L&M had the highest proportion of ENDS use among the non-premium bands. Oral nicotine/nicotine pouches were used by less than 5% of individuals for all brands in 2023 except American Spirit, with 7.1% of American Spirit users reporting oral nicotine/nicotine pouch use.

For other substances, alcohol use was greatest among those who reported smoking American Spirit, followed by Camel, and exhibited moderate to substantial decreases between 2013-2023. Past 30-day marijuana use was greatest among those who reported smoking American Spirit in 2013 and 2023, followed by Newport and Camel.

**Supplemental Table 1. Analysis sample by wave**

| Sample | W1 | W2 | W3 | W4 | W5 | W6 | W7 | Total^b^ observations across time |
| --- | --- | --- | --- | --- | --- | --- | --- | --- |
|  | W1 cohort^a^ | | | W4 cohort | | | W7 cohort |  |
|  | 2013/14 | 2014/15 | 2015/16 | 2016/17 | 2018/19 | 2020 | 2022/23 |  |
| Youth (12 to 17 yo) | 450 | 333 | 209 | 311 | 192 | 36 | 69 | 1,600 |
| Adults (18 + yo) | 11,040 | 9,401 | 9,564 | 10,613 | 8,541 | 5,868 | 5,391 | 60,418 |
| Total participants | 11,490 | 9,734 | 9,773 | 10,924 | 8,733 | 5,904 | 5,460 |  |
| Notes: W: Wave; yo: years old. | | | |  |  |  |  |  |

Notes. ^a^The W1 Cohort was followed over time, and replenished at W4 and W7, resulting in the W4 and W7 Cohorts, respectively. See Restricted Use Files User Guide for more information.

^b^Eligible analysis sample included a total of 62,018 observations across time among 19,722 unique individuals (1,201 youth and18,521 adults) for the study.

**Supplemental Table 2. List of premium and non-premium cigarette brands used in Wave 1-Wave 7 of the PATH Study**

| **Premium** | **Non-Premium** |
| --- | --- |
| American Spirit  Benson & Hedges  Camel  Capri  Carlton  Davidoff  Dunhill  Kent  Kool  Lucky Strike  Marlboro  Merit  More  Nat Sherman  Newport  Parliament  Players  Salem  Saratoga  Tareyton  Virginia Slims  Winston | 1839  305's  Basic  BLK  Bronson  Bugler  Carnival  Cheyenne  Criss Cross  Crown  Decade  Doral  Double Diamond  Eagle  Edgefield  Eve  Exeter  Fortuna  Gambler  Golden Harvest  Good Times  GPC  Grand Prix  Kentucky Select  Kentucky's Best  King Mountain  L & M  Liggett Select  Maverick  Middleton's  Misty  Monarch  Montclair  Mustang  Native  Nat’s  Old Gold  Palermo  Pall Mall  Pyramid  Rave  Remington  Santa Fe  Seneca  Shield  Signal  Smoker’s Choice  Sonoma  Southern Steel  Tahoe  Talon  The Good Stuff  Timeless Time  Top  Tourney  USA Gold  Wave  Wildhorse  Prime Time  Some other brand |

Note: Initial premium brands were selected from Cornelius et al.^5^ Expert reviews from authors (MR, RO, CD) provided feedback for brands not included in that paper, as well as re-classifying L&M and Eve into the non-premium category based on brand descriptions from parent company reports.^31,32^ The following were provided as cigarette brands but are not and were excluded from analysis: Al Capone, Black & Mild, BlackStone, Blu Cigs, Clipper, Djarum, NJOY, Phillies, Swisher Sweets, MarkTen, Vuse, Juul.

**Supplemental Table 3**. **Variable and measures definitions**

| **Variables** | **Definitions** | **Notes** |
| --- | --- | --- |
| **Variables used to define analysis sample at each wave** | | |
| Age^a^ | 12 years and older | --- |
| P30D Cigarette Use | Past 30-Day use of cigarettes | --- |
| Manufactured cigarette brand | Manufactured cigarette brands, e.g.,  1125 = Marlboro  1144 = Newport  1152 = Pall Mall  1033 = Camel | Respondents who were asked the brand question at Wave (W)1 and W2 or knew the name of the brand of manufactured cigarettes they usually smoke at W3 through W7.  At W1 the PATH Study asked adults: “What brand of cigarettes do (did) you usually (last) smoke?”  And asked youth “What brand of cigarette do (did) you usually (last) smoke?” (ASK: Past 30 day “Not Light” users)  At W7 the PATH Study asked adults: “What brand of cigarettes do (did) you usually (last) smoke?” (ASK: Adult respondents who are current manufactured cigarette smokers or recent former cigarette smokers and know the name of the brand of manufactured cigarettes they usually smoke or last smoked.)  And asked youth respondents who are past 30-day cigarette smokers who know the name of the brand of cigarettes they usually smoke or last smoked, “What brand of cigarette do (did) you usually (last) smoke?”  Used AC1048MC_BRAND for adults and YC1048_BRAND for youth. |
| P30D Cigarette menthol/mint flavor | Menthol or mint | In past 30 days, smoked cigarettes flavored to taste like menthol or mint.  PATH questions were asked slightly differently from W1 compared to W2 and later waves with the development/evolvement of instruments. Specifically, we used R01_AC1050MC and R02_AC1131_01 for adults and R01_YC9108 and R02_YC1131_01 for youth at W1 and W2. AC1130MC for adults and YC1130 for youth at W3 through W7. |
| P30D Cigarette consumption | 1 to 3000 | Total number of cigarettes consumed in the past 30 days, which was computed by multiplying the average number of cigarettes consumed each day and the total number of days that respondents reported smoking in the past 30 days. Specifically, we used the following variables to derive the P30D cigarette consumption measure for adults at each wave:  AC1022: In past 30 days, number of days smoked cigarettes  AC1021_NN: Average number of cigarettes now smoked each day - Number  AC1021_UN: Average number of cigarettes now smoked each day - Unit  AC1023_NN: In past 30 days, average number of cigarettes smoked per day on days smoked - Number  AC1023_UN: In past 30 days, average number of cigarettes smoked per day on days smoked - Unit  For youth we used two variables:  YC1022: In past 30 days, number of days smoked cigarettes  YC1023: In past 30 days, number of cigarettes smoked per day on the days that you smoked.  Special notes:   - This measure was derived to use for (1) adjusting BRR weights (each of the BRR weights was adjusted by multiplying this measure) for both youth and adults, and (2) computing P30D Cigarettes Smoked Per Day for adults. - Those who reported above 5 packs/day (100 CPD or 3000 cigarettes during P30D) accounted for <1% of the sample. We recoded >5 packs/day to 5 to address outliers (i.e., trimmed >3000 cigarettes during P30D to 3000). - Those who reported 0 cigarettes during P30D accounted for <=1% of the sample at each wave, we recoded them to 1 to prevent them from being dropped (i.e., standard BRR weighting method, since no weight adjustment was used because P30D Cigarette consumption is 1). - For youth at each wave, the response options for the question of “In the past 30 days, on the days you smoked, how many cigarettes did you smoke per day? A pack usually has 20 cigarettes in it.” were ordinal with range values. We recoded accordingly to be comparable to that of adults, using CPD as unit:   1 = Less than 1 per day (rounded up to 1 CPD, after examining distributions of the measure of P30D Cigarette consumption)  2 = 1 per day (recoded to 1)  3 = 2 to 5 per day (recoded to 3.5)  4 = 6 to 10 per day (recoded to 8)  5 = 11 to 20 per day (recoded to 15.5)  6 = More than 20 per day (recoded to 25) |
| **Wave (W) 1 - W7 variables** | | |
| Age^a^ | 1 = 12 to 17 years old  2 = 18 to 24 years old  3 = 25 to 54 years old  4 = 55+ years old | --- |
| Biological Sex^a^ | 1 = Male  2 = Female | --- |
| Race/ethnicity^a^ | 1 = Non-Hispanic White  2 = Non-Hispanic Black  3 = Hispanic  4 = Non-Hispanic Other Race (including Multiracial) | --- |
| Household Income | 1 = Less than $10,000  2 = $10,000 to $24,999  3 = $25,000 to $49,999  4 = $50,000 to $99,999  5 = $100,000 or more  6 = Not Reported | The PATH Study only collected household income data for adults at W1, and both adults and youth at W2 and later waves. |
| P30D Daily/Nondaily Cigarette Smoking | 1 = Yes  2 = No | --- |
| P30D E-Cigarette Use | 1 = Yes  2 = No | The PATH Study asked about “e-cigarettes” at W1 and “e-products” (i.e., e-cigarettes, e-cigars, e-pipes, and e-hookah) at W2 and later waves. |
| P30D Other combustible | 1 = Yes  2 = No | Other combustible products includes traditional cigars, cigarillos, filtered cigars, pipe tobacco, and hookah tobacco. Used any of these products was coded as a “Yes”. |
| P30D Smokeless tobacco | 1 = Yes  2 = No | Smokeless tobacco includes smokeless tobacco and snus. Used either of these products was coded as a “Yes”. |
| P30D Oral Nicotine/Nicotine Pouches (W7 only) | 1 = Yes  2 = No | Oral nicotine includes lozenges, discs, tablets, gum, toothpicks, dissolvable tobacco, and related products, but exclude nicotine replacement therapy. Nicotine pouches were asked about separately. Used either of these products was coded as a “Yes”. |
| P30D Alcohol use | 1 = Yes  2 = No | Questions about P30D alcohol use were asked differently at W1; for example, the PATH Study asked, "How long has it been since you last used alcohol?" at W1, and an endorsement of “Within the past 30 days” was coded as “Yes”. Whereas at W7 asked directly, " Have you used alcohol within the past 30 days?". |
| P30D Marijuana use | 1 = Yes  2 = No | Questions about P30D marijuana use were asked differently at W1; for example, the PATH Study asked, " How long has it been since you last used marijuana, hash, THC, grass, pot, or weed?" at W1, and an endorsement of “Within the past 30 days” was coded as “Yes”. Whereas at W7 asked directly, "Have you used marijuana within the past 30 days?". |
| Mental Health (P30D GAIN internalizing scale) | 1 = No/Low  2 = Moderate  3 = High | The PATH Study asked “When was the last time that you had significant problems with”:   - Feeling very trapped, lonely, sad, blue, depressed or hopeless about the future - Sleep trouble - such as bad dreams, sleeping restlessly or falling asleep during the day - Feeling very anxious, nervous, tense, scared, panicked or like something bad was going to happen - Becoming very distressed and upset when something reminded you of the past   An endorsement of “Past month” was recoded as 1 (other valid data were recoded as 0), and summary scores ranged from 0 to 4 for internalizing problems. Respondents were categorized into no/low (0–1 symptoms), moderate (2–3 symptoms), or high 4 symptoms for the scale, consistent with Conway et al. 2017. |

Notes: ^a^Missing data on age, biological sex, race, and Hispanic ethnicity were imputed at W1, and sex, race, and Hispanic ethnicity were also imputed at W4 and W7 as described in the PATH Study Public Use Files User Guide.^30^

**Supplemental Table 4. Weights used to maximize the sample size at each wave**

| Wave 1 | W1 Cross-sectional weights for the W1 Cohort |
| --- | --- |
| Wave 2 | W2 Single-wave longitudinal weights for the W1 Cohort |
| Wave 3 | W3 Single-wave longitudinal weights for the W1 Cohort |
| Wave 4 | W4 Cross-sectional weights for the W4 cohort |
| Wave 5 | W5 Single-wave longitudinal weights for the W4 Cohort |
| Wave 6 | W6 Single-wave weights for the W4 Cohort |
| Wave 7 | W7 Cross-sectional weights for the W7 Cohort |

Notes: A full sample and 100 Balance Repeated Replicate (BRR) weights at each wave were used in analyses. Weighted estimates from Waves 1-7 of the PATH Study represent the resident population of the U.S. ages 12 years and older at the time of data collection who were part of the U.S. civilian, noninstitutionalized population at W1, W4 or W7, for each cohort, respectively.

**Supplemental Table 5. Demographic and substance use profile of smokers of the top 10 brands, 2013/14 and 2022/23**

|  | **Top 1 Brand**  **(Marlboro)** | | **Top 2 Brand** **(Newport)** | | **Top 3 Brand**  **(Camel)** | | Top 4 Brand  (Pall Mall) | | Top 5 Brand  (Maverick) | |
| --- | --- | --- | --- | --- | --- | --- | --- | --- | --- | --- |
|  | W1 (2013/14) | W7 (2022/23) | W1 (2013/14) | W7 (2022/23) | W1 (2013/14) | W7 (2022/23) | W1 (2013/14) | W7 (2022/23) | W1 (2013/14) | W7 (2022/23) |
|  | Weighted % (95%CI) | Weighted % (95%CI) | Weighted % (95%CI) | Weighted % (95%CI) | Weighted % (95%CI) | Weighted % (95%CI) | Weighted % (95%CI) | Weighted % (95%CI) | Weighted % (95%CI) | Weighted % (95%CI) |
| Age |  |  |  |  |  |  |  |  |  |  |
| 12 to 17 years old | 2.8 ( 2.3, 3.3) | 0.7 ( 0.4, 1.2) | 1.6 ( 1.2, 2.3) | 0.4 ( 0.2, 1.0) | 2.8 ( 2.0, 3.9) | 0.9 ( 0.4, 2.1) | 0.6 ( 0.2, 1.5) | 0.4 ( 0.1, 2.2) | 1.7 ( 0.7, 4.2) | 0.0 NA |
| 18 to 24 | 16.3 (15.1, 17.6) | 6.4 ( 5.3, 7.7) | 18.5 (16.8, 20.5) | 5.0 ( 3.8, 6.4) | 25.7 (23.3, 28.2) | 10.0 ( 7.8, 12.8) | 3.9 ( 2.7, 5.6) | 0.8 ( 0.2, 2.8) | 7.9 ( 5.2, 11.8) | 2.1 ( 0.7, 6.1) |
| 25-54 | 66.1 (64.5, 67.6) | 67.5 (63.8, 71.1) | 68.3 (65.9, 70.6) | 70.4 (66.6, 74.0) | 64.4 (61.7, 67.0) | 77.7 (73.2, 81.6) | 57.0 (52.9, 60.9) | 46.3 (38.7, 54.1) | 63.5 (57.0, 69.5) | 43.1 (34.3, 52.4) |
| 55 + | 14.9 (13.6, 16.3) | 25.4 (22.2, 28.9) | 11.5 ( 9.8, 13.5) | 24.2 (20.7, 28.1) | 7.1 ( 5.6, 9.0) | 11.5 ( 8.4, 15.4) | 38.5 (34.3, 43.0) | 52.6 (44.9, 60.1) | 26.9 (21.2, 33.5) | 54.8 (45.3, 64.0) |
| Biological Sex |  |  |  |  |  |  |  |  |  |  |
| Male | 57.7 (56.3, 59.2) | 53.2 (49.9, 56.4) | 54.8 (52.3, 57.3) | 54.6 (50.6, 58.6) | 59.7 (56.2, 63.1) | 60.2 (54.7, 65.3) | 52.4 (48.1, 56.7) | 50.0 (43.1, 57.0) | 55.9 (49.8, 61.8) | 50.9 (40.2, 61.5) |
| Female | 42.3 (40.8, 43.7) | 46.8 (43.6, 50.1) | 45.2 (42.7, 47.7) | 45.4 (41.4, 49.4) | 40.3 (36.9, 43.8) | 39.8 (34.7, 45.3) | 47.6 (43.3, 51.9) | 50.0 (43.0, 56.9) | 44.1 (38.2, 50.2) | 49.1 (38.5, 59.8) |
| Race/Ethnicity |  |  |  |  |  |  |  |  |  |  |
| White NH | 77.7 (76.0, 79.2) | 71.3 (68.8, 73.7) | 29.2 (26.4, 32.1) | 24.4 (21.3, 27.7) | 76.3 (73.7, 78.8) | 62.9 (57.5, 68.1) | 83.7 (80.3, 86.7) | 79.7 (72.6, 85.3) | 55.3 (47.9, 62.4) | 52.4 (42.4, 62.3) |
| Black NH | 2.4 ( 1.9, 3.0) | 2.7 ( 1.9, 3.6) | 50.3 (47.1, 53.5) | 49.7 (45.5, 53.9) | 2.9 ( 2.1, 4.1) | 4.6 ( 3.1, 6.9) | 6.4 ( 4.5, 8.9) | 5.0 ( 2.9, 8.6) | 34.1 (27.0, 42.0) | 31.7 (23.3, 41.4) |
| Hispanic | 13.0 (11.9, 14.3) | 17.4 (15.2, 19.8) | 15.9 (13.7, 18.4) | 18.6 (15.2, 22.6) | 13.7 (11.7, 16.0) | 23.9 (19.0, 29.7) | 6.1 ( 4.4, 8.6) | 7.3 ( 4.1, 12.6) | 5.9 ( 3.6, 9.4) | 6.7 ( 3.0, 14.1) |
| Other | 6.9 ( 6.0, 7.9) | 8.7 ( 7.1, 10.6) | 4.7 ( 3.7, 5.9) | 7.3 ( 5.6, 9.4) | 7.0 ( 5.7, 8.6) | 8.5 ( 5.6, 12.7) | 3.8 ( 2.6, 5.5) | 7.9 ( 4.3, 14.3) | 4.7 ( 2.6, 8.4) | 9.2 ( 3.6, 21.6) |
| Sexual Orientation^a^ |  |  |  |  |  |  |  |  |  |  |
| Lesbian, Gay, Bisexual or Something Else | 7.0 ( 6.2, 7.9) | 11.2 ( 9.6, 13.1) | 8.7 ( 7.4, 10.2) | 13.9 (11.8, 16.4) | 8.6 ( 7.0, 10.5) | 15.7 (12.2, 20.0) | 6.8 ( 5.1, 8.9) | 6.8 ( 3.7, 12.4) | 8.1 ( 5.1, 12.6) | 12.0 ( 7.5, 18.8) |
| Straight | 93.0 (92.1, 93.8) | 88.8 (86.9, 90.4) | 91.3 (89.8, 92.6) | 86.1 (83.6, 88.2) | 91.4 (89.5, 93.0) | 84.3 (80.0, 87.8) | 93.2 (91.1, 94.9) | 93.2 (87.6, 96.3) | 91.9 (87.4, 94.9) | 88.0 (81.2, 92.5) |
| Household Income |  |  |  |  |  |  |  |  |  |  |
| Less than $10,000 | 15.5 (14.2, 16.9) | 10.4 ( 8.8, 12.3) | 29.4 (26.9, 32.1) | 28.5 (24.9, 32.4) | 16.5 (14.0, 19.4) | 19.4 (15.9, 23.4) | 17.2 (14.1, 20.8) | 12.3 ( 8.4, 17.6) | 31.2 (25.3, 37.8) | 22.3 (15.7, 30.9) |
| $10,000 to $24,999 | 21.7 (20.1, 23.5) | 17.3 (15.4, 19.4) | 28.4 (25.8, 31.2) | 20.4 (17.3, 23.9) | 24.4 (21.8, 27.3) | 13.9 (10.6, 17.9) | 31.9 (28.3, 35.6) | 24.2 (17.4, 32.7) | 32.6 (25.9, 40.0) | 29.7 (20.8, 40.4) |
| $25,000 to $49,999 | 23.8 (22.3, 25.2) | 23.8 (21.1, 26.8) | 20.4 (18.1, 22.8) | 25.6 (22.3, 29.1) | 26.7 (24.0, 29.7) | 23.6 (19.8, 27.9) | 27.5 (23.3, 32.1) | 22.3 (16.9, 28.8) | 17.4 (12.9, 22.9) | 21.9 (13.3, 33.7) |
| $50,000 to $99,999 | 20.3 (18.9, 21.8) | 26.9 (24.6, 29.3) | 10.7 ( 8.9, 12.7) | 16.7 (14.1, 19.7) | 19.2 (16.8, 21.8) | 25.6 (20.8, 31.0) | 14.3 (11.2, 18.2) | 24.3 (17.6, 32.6) | 8.3 ( 5.5, 12.4) | 16.4 ( 9.2, 27.7) |
| $100,000 or more | 10.0 ( 8.8, 11.3) | 16.9 (14.6, 19.4) | 3.7 ( 2.6, 5.2) | 4.6 ( 3.2, 6.4) | 7.6 ( 5.9, 9.9) | 15.1 (11.5, 19.6) | 2.1 ( 1.2, 3.6) | 8.7 ( 5.1, 14.3) | 2.0 ( 0.8, 5.1) | 4.3 ( 1.8, 9.7) |
| Not Reported | 8.7 ( 7.8, 9.7) | 4.7 ( 3.7, 6.0) | 7.5 ( 6.0, 9.3) | 4.2 ( 2.7, 6.7) | 5.5 ( 4.4, 7.0) | 2.5 ( 1.5, 4.2) | 7.0 ( 5.2, 9.3) | 8.2 ( 5.0, 13.1) | 8.5 ( 5.4, 13.3) | 5.5 ( 2.0, 13.7) |
| P30D Daily/Nondaily Cigarette Smoking |  |  |  |  |  |  |  |  |  |  |
| Yes | 79.1 (77.6, 80.6) | 72.3 (69.7, 74.9) | 82.8 (80.2, 85.1) | 72.1 (69.1, 75.0) | 68.0 (65.1, 70.8) | 62.8 (57.0, 68.3) | 92.4 (90.2, 94.2) | 90.0 (84.5, 93.6) | 93.1 (89.4, 95.6) | 86.0 (79.1, 90.8) |
| No | 20.9 (19.4, 22.4) | 27.7 (25.1, 30.3) | 17.2 (14.9, 19.8) | 27.9 (25.0, 30.9) | 32.0 (29.2, 34.9) | 37.2 (31.7, 43.0) | 7.6 ( 5.8, 9.8) | 10.0 ( 6.4, 15.5) | 6.9 ( 4.4, 10.6) | 14.0 ( 9.2, 20.9) |
| P30D ENDS Use^b^ |  |  |  |  |  |  |  |  |  |  |
| Yes | 26.0 (24.4, 27.5) | 26.0 (23.9, 28.2) | 21.9 (19.6, 24.4) | 22.4 (19.2, 26.0) | 32.1 (29.4, 35.0) | 32.5 (27.0, 38.6) | 20.7 (17.8, 23.9) | 14.1 ( 9.6, 20.3) | 20.2 (15.7, 25.5) | 18.9 (12.9, 26.7) |
| No | 74.0 (72.5, 75.6) | 74.0 (71.8, 76.1) | 78.1 (75.6, 80.4) | 77.6 (74.0, 80.8) | 67.9 (65.0, 70.6) | 67.5 (61.4, 73.0) | 79.3 (76.1, 82.2) | 85.9 (79.7, 90.4) | 79.8 (74.5, 84.3) | 81.1 (73.3, 87.1) |
| P30D Other Combustible^c^ |  |  |  |  |  |  |  |  |  |  |
| Yes | 21.1 (19.6, 22.6) | 11.6 ( 9.9, 13.4) | 34.7 (32.2, 37.2) | 26.9 (23.7, 30.4) | 27.3 (24.8, 29.8) | 20.2 (15.9, 25.2) | 17.1 (14.3, 20.3) | 7.3 ( 4.4, 11.8) | 30.5 (24.5, 37.3) | 18.5 (12.5, 26.4) |
| No | 78.9 (77.4, 80.4) | 88.4 (86.6, 90.1) | 65.3 (62.8, 67.8) | 73.1 (69.6, 76.3) | 72.7 (70.2, 75.2) | 79.8 (74.8, 84.1) | 82.9 (79.7, 85.7) | 92.7 (88.2, 95.6) | 69.5 (62.7, 75.5) | 81.5 (73.6, 87.5) |
| P30D Smokeless Tobacco^c^ |  |  |  |  |  |  |  |  |  |  |
| Yes | 9.0 ( 8.2, 9.9) | 4.9 ( 3.6, 6.5) | 3.8 ( 2.9, 4.8) | 1.6 ( 1.0, 2.6) | 10.0 ( 8.3, 11.9) | 5.2 ( 3.5, 7.8) | 4.8 ( 3.2, 7.3) | 3.6 ( 1.7, 7.7) | 8.1 ( 5.2, 12.3) | 0.6 ( 0.1, 3.8) |
| No | 91.0 (90.1, 91.8) | 95.1 (93.5, 96.4) | 96.2 (95.2, 97.1) | 98.4 (97.4, 99.0) | 90.0 (88.1, 91.7) | 94.8 (92.2, 96.5) | 95.2 (92.7, 96.8) | 96.4 (92.3, 98.3) | 91.9 (87.7, 94.8) | 99.4 (96.2, 99.9) |
| P30D Oral Nicotine/Nicotine Pouches^c^ |  |  |  |  |  |  |  |  |  |  |
| Yes | N/A | 3.7 ( 2.7, 5.0) | N/A | 1.6 ( 0.9, 2.9) | N/A | 3.9 ( 2.5, 6.1) | N/A | 4.9 ( 2.2, 10.8) | N/A | 1.6 ( 0.4, 6.1) |
| No | N/A | 96.3 (95.0, 97.3) | N/A | 98.4 (97.1, 99.1) | N/A | 96.1 (93.9, 97.5) | N/A | 95.1 (89.2, 97.8) | N/A | 98.4 (93.9, 99.6) |
| P30D Alcohol Use |  |  |  |  |  |  |  |  |  |  |
| Yes | 59.8 (57.9, 61.6) | 54.4 (51.0, 57.8) | 59.9 (57.0, 62.8) | 47.4 (43.8, 51.0) | 70.0 (67.1, 72.7) | 57.3 (51.7, 62.8) | 53.1 (48.6, 57.5) | 51.8 (44.4, 59.0) | 52.4 (44.7, 59.9) | 48.1 (36.5, 59.8) |
| No | 40.2 (38.4, 42.1) | 45.6 (42.2, 49.0) | 40.1 (37.2, 43.0) | 52.6 (49.0, 56.2) | 30.0 (27.3, 32.9) | 42.7 (37.2, 48.3) | 46.9 (42.5, 51.4) | 48.2 (41.0, 55.6) | 47.6 (40.1, 55.3) | 51.9 (40.2, 63.5) |
| P30D Marijuana Use |  |  |  |  |  |  |  |  |  |  |
| Yes | 18.7 (17.4, 20.1) | 33.4 (31.0, 35.9) | 27.4 (24.9, 30.0) | 43.1 (39.3, 46.9) | 29.3 (26.5, 32.2) | 42.7 (37.6, 47.9) | 13.9 (11.5, 16.7) | 29.3 (22.5, 37.2) | 18.8 (14.3, 24.4) | 25.7 (18.3, 34.9) |
| No | 81.3 (79.9, 82.6) | 66.6 (64.1, 69.0) | 72.6 (70.0, 75.1) | 56.9 (53.1, 60.7) | 70.7 (67.8, 73.5) | 57.3 (52.1, 62.4) | 86.1 (83.3, 88.5) | 70.7 (62.8, 77.5) | 81.2 (75.6, 85.7) | 74.3 (65.1, 81.7) |
| Mental Health (P30D GAIN internalizing scale) |  |  |  |  |  |  |  |  |  |  |
| Low | 71.4 (69.9, 72.9) | 73.6 (71.0, 76.0) | 69.7 (67.1, 72.1) | 71.8 (68.2, 75.2) | 68.4 (64.7, 71.9) | 65.8 (60.3, 71.0) | 66.9 (63.0, 70.5) | 75.5 (68.2, 81.5) | 63.7 (57.6, 69.3) | 75.0 (65.6, 82.5) |
| Moderate | 17.9 (16.8, 19.1) | 17.2 (15.1, 19.6) | 18.1 (16.3, 20.1) | 15.7 (13.3, 18.3) | 19.5 (16.9, 22.3) | 19.9 (16.5, 23.8) | 19.8 (17.0, 23.0) | 17.0 (12.2, 23.3) | 23.6 (18.9, 29.2) | 8.8 ( 5.1, 14.8) |
| High | 10.6 ( 9.6, 11.7) | 9.2 ( 7.8, 10.7) | 12.2 (10.6, 14.0) | 12.5 ( 9.8, 15.9) | 12.1 (10.1, 14.4) | 14.3 (10.7, 18.7) | 13.3 (10.4, 16.8) | 7.5 ( 4.5, 12.3) | 12.7 ( 9.2, 17.3) | 16.2 (10.4, 24.5) |
| P30D Cigarettes Smoked Per Day |  |  |  |  |  |  |  |  |  |  |
| Mean^d^ | 13.9 (13.5, 14.3) | 12.1 (11.5, 12.7) | 13.0 (12.3, 13.8) | 9.9 ( 9.1, 10.7) | 11.1 (10.5, 11.7) | 10.2 ( 8.9, 11.6) | 17.1 (16.0, 18.3) | 16.9 (14.8, 19.0) | 17.1 (14.7, 19.6) | 14.8 (12.7, 17.0) |
| **Supplemental Table 5 continued**  **(Brands 6-10)** | Top 6 Brand  (L & M) | | Top 7 Brand  (Sonoma) | | Top 8 Brand  (305's) | | **Top 9 Brand**  **(Kool)** | | **Top10 Brand** **(American Spirit)** | |
|  | W1 (2013/14) | W7 (2022/23) | W1 (2013/14) | W7 (2022/23) | W1 (2013/14) | W7 (2022/23) | W1 (2013/14) | W7 (2022/23) | W1 (2013/14) | W7 (2022/23) |
|  | Weighted % (95%CI) | Weighted % (95%CI) | Weighted % (95%CI) | Weighted % (95%CI) | Weighted % (95%CI) | Weighted % (95%CI) | Weighted % (95%CI) | Weighted % (95%CI) | Weighted % (95%CI) | Weighted % (95%CI) |
| Age |  |  |  |  |  |  |  |  |  |  |
| 12 to 17 years old | 0.6 ( 0.1, 2.5) | 0.2 ( 0.0, 3.2) | 1.0 ( 0.2, 6.1) | 0.0 NA | 0.8 ( 0.1, 5.4) | 0.0 NA | 1.4 ( 0.6, 3.5) | 0.0 NA | 2.6 ( 1.2, 5.4) | 0.4 ( 0.1, 2.1) |
| 18 to 24 | 20.3 (15.8, 25.6) | 5.6 ( 2.8, 11.0) | 5.2 ( 2.2, 12.2) | 6.3 ( 2.9, 13.3) | 1.9 ( 0.5, 7.1) | 2.9 ( 0.9, 9.4) | 7.8 ( 5.3, 11.4) | 2.5 ( 0.9, 6.7) | 18.8 (14.3, 24.5) | 14.2 (10.5, 19.0) |
| 25-54 | 62.0 (54.1, 69.2) | 72.0 (62.0, 80.2) | 59.7 (49.0, 69.4) | 54.1 (41.3, 66.5) | 62.1 (51.5, 71.6) | 49.9 (38.5, 61.4) | 55.1 (46.8, 63.2) | 45.2 (32.4, 58.6) | 60.1 (52.1, 67.6) | 61.2 (53.9, 68.1) |
| 55 + | 17.2 (12.2, 23.7) | 22.2 (14.9, 31.8) | 34.1 (24.9, 44.7) | 39.5 (28.1, 52.1) | 35.2 (26.3, 45.3) | 47.2 (35.0, 59.6) | 35.6 (27.6, 44.6) | 52.4 (38.2, 66.2) | 18.4 (13.1, 25.2) | 24.2 (18.1, 31.5) |
| Biological Sex |  |  |  |  |  |  |  |  |  |  |
| Male | 50.2 (43.0, 57.4) | 50.9 (41.0, 60.8) | 45.7 (35.0, 56.8) | 38.6 (27.7, 50.7) | 46.8 (34.6, 59.5) | 46.4 (33.0, 60.4) | 63.5 (57.9, 68.8) | 65.4 (53.2, 75.9) | 60.9 (54.6, 66.9) | 63.7 (56.7, 70.1) |
| Female | 49.8 (42.6, 57.0) | 49.1 (39.2, 59.0) | 54.3 (43.2, 65.0) | 61.4 (49.3, 72.3) | 53.2 (40.5, 65.4) | 53.6 (39.6, 67.0) | 36.5 (31.2, 42.1) | 34.6 (24.1, 46.8) | 39.1 (33.1, 45.4) | 36.3 (29.9, 43.3) |
| Race/Ethnicity |  |  |  |  |  |  |  |  |  |  |
| White NH | 90.9 (86.8, 93.9) | 77.1 (69.2, 83.5) | 86.4 (71.2, 94.3) | 81.3 (70.5, 88.8) | 82.8 (67.5, 91.8) | 83.9 (71.4, 91.6) | 32.0 (25.0, 40.0) | 35.4 (23.1, 49.9) | 79.3 (73.1, 84.4) | 78.5 (71.8, 83.9) |
| Black NH | 0.6 ( 0.2, 2.6) | 10.0 ( 6.0, 16.4) | 7.4 ( 2.2, 21.7) | 6.9 ( 2.5, 17.5) | 7.1 ( 2.2, 20.4) | 3.2 ( 1.0, 9.7) | 51.1 (41.4, 60.7) | 44.5 (33.9, 55.6) | 2.4 ( 1.0, 5.9) | 3.3 ( 1.5, 7.4) |
| Hispanic | 3.2 ( 1.6, 6.1) | 4.0 ( 1.8, 8.9) | 4.7 ( 1.8, 11.8) | 1.3 ( 0.3, 6.3) | 6.3 ( 2.1, 17.7) | 11.6 ( 5.3, 23.5) | 11.1 ( 7.4, 16.1) | 7.4 ( 3.6, 14.7) | 9.6 ( 6.5, 13.9) | 8.6 ( 5.8, 12.6) |
| Other | 5.2 ( 3.1, 8.7) | 8.8 ( 4.1, 18.1) | 1.5 ( 0.3, 7.0) | 10.5 ( 5.7, 18.4) | 3.8 ( 1.4, 10.1) | 1.3 ( 0.2, 7.0) | 5.8 ( 2.4, 13.5) | 12.7 ( 5.3, 27.5) | 8.6 ( 5.0, 14.4) | 9.6 ( 5.4, 16.4) |
| Sexual Orientation^a^ |  |  |  |  |  |  |  |  |  |  |
| Lesbian, Gay, Bisexual or Something Else | 6.2 ( 3.6, 10.3) | 18.7 (12.2, 27.7) | 1.1 ( 0.2, 6.3) | 7.4 ( 3.6, 14.7) | 13.0 ( 6.6, 23.8) | 16.3 ( 9.7, 26.2) | 7.2 ( 4.5, 11.5) | 10.0 ( 5.9, 16.4) | 15.6 (10.3, 22.9) | 22.6 (16.8, 29.8) |
| Straight | 93.8 (89.7, 96.4) | 81.3 (72.3, 87.8) | 98.9 (93.7, 99.8) | 92.6 (85.3, 96.4) | 87.0 (76.2, 93.4) | 83.7 (73.8, 90.3) | 92.8 (88.5, 95.5) | 90.0 (83.6, 94.1) | 84.4 (77.1, 89.7) | 77.4 (70.2, 83.2) |
| Household Income |  |  |  |  |  |  |  |  |  |  |
| Less than $10,000 | 17.8 (13.0, 23.8) | 15.2 ( 9.3, 23.8) | 26.0 (17.8, 36.3) | 26.7 (18.8, 36.6) | 21.9 (13.9, 32.8) | 24.6 (15.7, 36.4) | 24.2 (19.5, 29.5) | 28.2 (17.3, 42.4) | 11.1 ( 7.7, 15.7) | 5.3 ( 3.2, 8.9) |
| $10,000 to $24,999 | 32.2 (25.2, 40.1) | 16.0 (10.7, 23.3) | 32.1 (21.1, 45.6) | 29.9 (21.5, 39.9) | 22.2 (14.9, 31.7) | 35.5 (25.4, 47.2) | 29.9 (24.9, 35.4) | 12.7 ( 8.1, 19.4) | 20.1 (15.5, 25.7) | 18.0 (12.4, 25.3) |
| $25,000 to $49,999 | 22.5 (17.7, 28.2) | 39.7 (30.0, 50.3) | 26.4 (18.1, 36.8) | 25.4 (17.7, 35.2) | 24.7 (14.0, 39.7) | 17.0 (10.3, 26.9) | 21.0 (16.3, 26.5) | 23.9 (16.4, 33.4) | 28.6 (22.5, 35.5) | 23.5 (18.3, 29.6) |
| $50,000 to $99,999 | 14.0 ( 9.3, 20.5) | 20.7 (13.4, 30.7) | 8.3 ( 4.1, 16.2) | 12.2 ( 7.0, 20.5) | 10.9 ( 5.1, 21.8) | 12.3 ( 6.1, 23.0) | 15.0 (10.8, 20.4) | 20.1 (12.1, 31.5) | 19.8 (14.6, 26.3) | 28.6 (20.9, 37.8) |
| $100,000 or more | 4.5 ( 1.9, 10.2) | 5.4 ( 1.6, 16.6) | 0.0 NA | 0.9 ( 0.1, 5.8) | 4.7 ( 1.9, 11.3) | 6.8 ( 2.0, 20.5) | 1.4 ( 0.6, 3.6) | 3.6 ( 1.4, 9.5) | 12.4 ( 8.1, 18.4) | 21.0 (15.6, 27.7) |
| Not Reported | 9.1 ( 5.6, 14.2) | 3.0 ( 1.0, 8.7) | 7.2 ( 2.5, 18.8) | 4.8 ( 1.7, 12.7) | 15.5 ( 8.6, 26.4) | 3.8 ( 1.3, 10.6) | 8.6 ( 5.7, 12.6) | 11.5 ( 5.3, 23.0) | 8.1 ( 4.9, 13.0) | 3.6 ( 1.7, 7.2) |
| P30D Daily/Nondaily Cigarette Smoking |  |  |  |  |  |  |  |  |  |  |
| Yes | 93.5 (88.6, 96.3) | 83.9 (72.8, 91.0) | 84.7 (71.3, 92.5) | 96.2 (90.1, 98.6) | 88.8 (80.8, 93.7) | 90.3 (78.4, 96.0) | 81.2 (76.1, 85.5) | 79.9 (68.8, 87.7) | 64.4 (57.9, 70.5) | 52.7 (44.7, 60.6) |
| No | 6.5 ( 3.7, 11.4) | 16.1 ( 9.0, 27.2) | 15.3 ( 7.5, 28.7) | 3.8 ( 1.4, 9.9) | 11.2 ( 6.3, 19.2) | 9.7 ( 4.0, 21.6) | 18.8 (14.5, 23.9) | 20.1 (12.3, 31.2) | 35.6 (29.5, 42.1) | 47.3 (39.4, 55.3) |
| P30D ENDS Use^b^ |  |  |  |  |  |  |  |  |  |  |
| Yes | 27.3 (21.4, 34.1) | 26.8 (18.5, 37.1) | 18.0 (11.0, 27.9) | 21.5 (11.4, 36.9) | 24.4 (16.8, 34.0) | 24.3 (16.2, 34.9) | 16.7 (12.6, 21.8) | 17.7 ( 9.6, 30.5) | 31.4 (25.0, 38.7) | 32.4 (25.7, 39.8) |
| No | 72.7 (65.9, 78.6) | 73.2 (62.9, 81.5) | 82.0 (72.1, 89.0) | 78.5 (63.1, 88.6) | 75.6 (66.0, 83.2) | 75.7 (65.1, 83.8) | 83.3 (78.2, 87.4) | 82.3 (69.5, 90.4) | 68.6 (61.3, 75.0) | 67.6 (60.2, 74.3) |
| P30D Other Combustible^c^ |  |  |  |  |  |  |  |  |  |  |
| Yes | 18.2 (13.2, 24.6) | 19.0 (10.6, 31.7) | 19.3 ( 9.7, 34.7) | 10.2 ( 5.6, 18.1) | 29.8 (21.5, 39.7) | 18.0 ( 8.5, 34.2) | 24.6 (18.8, 31.5) | 24.3 (14.0, 38.7) | 23.9 (19.0, 29.7) | 15.3 (10.5, 21.7) |
| No | 81.8 (75.4, 86.8) | 81.0 (68.3, 89.4) | 80.7 (65.3, 90.3) | 89.8 (81.9, 94.4) | 70.2 (60.3, 78.5) | 82.0 (65.8, 91.5) | 75.4 (68.5, 81.2) | 75.7 (61.3, 86.0) | 76.1 (70.3, 81.0) | 84.7 (78.3, 89.5) |
| P30D Smokeless Tobacco^c^ |  |  |  |  |  |  |  |  |  |  |
| Yes | 7.6 ( 5.0, 11.6) | 9.9 ( 3.7, 23.8) | 11.7 ( 6.2, 21.1) | 8.6 ( 3.4, 20.2) | 1.3 ( 0.2, 6.1) | 0.0 NA | 3.8 ( 2.0, 7.3) | 4.5 ( 2.1, 9.4) | 6.0 ( 3.6, 9.9) | 4.0 ( 2.1, 7.7) |
| No | 92.4 (88.4, 95.0) | 90.1 (76.2, 96.3) | 88.3 (78.9, 93.8) | 91.4 (79.8, 96.6) | 98.7 (93.9, 99.8) | 100.0 NA | 96.2 (92.7, 98.0) | 95.5 (90.6, 97.9) | 94.0 (90.1, 96.4) | 96.0 (92.3, 97.9) |
| P30D Oral Nicotine/Nicotine Pouches^c^ |  |  |  |  |  |  |  |  |  |  |
| Yes | N/A | 4.9 ( 1.8, 12.4) | N/A | 4.1 ( 0.7, 20.7) | N/A | 1.6 ( 0.3, 7.4) | N/A | 1.4 ( 0.3, 5.7) | N/A | 7.1 ( 3.5, 13.6) |
| No | N/A | 95.1 (87.6, 98.2) | N/A | 95.9 (79.3, 99.3) | N/A | 98.4 (92.6, 99.7) | N/A | 98.6 (94.3, 99.7) | N/A | 92.9 (86.4, 96.5) |
| P30D Alcohol Use |  |  |  |  |  |  |  |  |  |  |
| Yes | 52.1 (44.5, 59.7) | 46.0 (34.9, 57.5) | 35.6 (21.8, 52.2) | 49.3 (32.4, 66.5) | 60.1 (47.4, 71.7) | 50.8 (35.6, 65.9) | 53.3 (45.5, 60.9) | 35.0 (25.1, 46.5) | 76.6 (70.3, 81.8) | 77.1 (69.1, 83.6) |
| No | 47.9 (40.3, 55.5) | 54.0 (42.5, 65.1) | 64.4 (47.8, 78.2) | 50.7 (33.5, 67.6) | 39.9 (28.3, 52.6) | 49.2 (34.1, 64.4) | 46.7 (39.1, 54.5) | 65.0 (53.5, 74.9) | 23.4 (18.2, 29.7) | 22.9 (16.4, 30.9) |
| P30D Marijuana Use |  |  |  |  |  |  |  |  |  |  |
| Yes | 19.4 (15.0, 24.7) | 41.3 (29.7, 53.8) | 10.8 ( 5.2, 21.3) | 35.8 (24.5, 48.9) | 14.7 ( 8.8, 23.5) | 40.4 (30.2, 51.5) | 18.3 (14.2, 23.2) | 40.1 (28.2, 53.5) | 36.2 (29.4, 43.6) | 49.6 (41.3, 57.8) |
| No | 80.6 (75.3, 85.0) | 58.7 (46.2, 70.3) | 89.2 (78.7, 94.8) | 64.2 (51.1, 75.5) | 85.3 (76.5, 91.2) | 59.6 (48.5, 69.8) | 81.7 (76.8, 85.8) | 59.9 (46.5, 71.8) | 63.8 (56.4, 70.6) | 50.4 (42.2, 58.7) |
| Mental Health (P30D GAIN internalizing scale) |  |  |  |  |  |  |  |  |  |  |
| Low | 62.6 (55.8, 69.0) | 74.1 (63.1, 82.7) | 67.9 (57.3, 76.9) | 62.4 (52.1, 71.7) | 72.9 (61.7, 81.8) | 67.1 (56.0, 76.6) | 72.1 (66.6, 77.1) | 85.7 (78.5, 90.7) | 63.0 (56.4, 69.1) | 66.1 (58.3, 73.0) |
| Moderate | 25.6 (20.1, 31.9) | 11.1 ( 6.8, 17.7) | 16.4 (10.0, 25.8) | 18.7 (11.6, 28.6) | 16.9 (10.6, 25.9) | 18.0 (11.0, 28.1) | 17.7 (13.6, 22.8) | 10.3 ( 5.6, 18.1) | 19.9 (15.1, 25.8) | 22.9 (16.8, 30.5) |
| High | 11.8 ( 7.7, 17.7) | 14.8 ( 8.0, 25.8) | 15.7 ( 9.5, 25.0) | 18.9 (12.2, 28.2) | 10.2 ( 5.5, 18.1) | 14.8 ( 8.6, 24.4) | 10.2 ( 7.0, 14.5) | 4.1 ( 1.8, 8.9) | 17.1 (12.7, 22.5) | 11.0 ( 7.6, 15.7) |
| P30D Cigarettes Smoked Per Day |  |  |  |  |  |  |  |  |  |  |
| Mean^d^ | 17.5 (16.2, 18.9) | 13.6 (11.2, 16.0) | 17.2 (14.8, 19.6) | 19.5 (12.6, 26.4) | 17.1 (15.1, 19.2) | 17.2 (13.4, 21.0) | 15.3 (13.0, 17.5) | 13.2 (11.4, 14.9) | 9.2 ( 7.9, 10.5) | 6.4 ( 5.5, 7.3) |

Notes: Abbreviations W1: Wave 1, W7: Wave 7; NH: Non-Hispanic; P30D: Past 30 days, ENDS: Electronic Nicotine Delivery Systems; GAIN: Global Appraisal of Needs. Estimates were weighted by W1 Cross-sectional weights for the W1 Cohort and W7 Cross-sectional weights for the W7 Cohort, respectively for W1 (2013/14) and W7(2022/23). Weighted percentages with Wilson confidence limits were presented. The Top 10 brands were ordered by Wave 7, and **bolded are premium brands**. ^a^The PATH Study PUF provides a 2-level sexual orientation variable for adults and exclude all sexual orientation variables for youth. ^b^The PATH Study asked about “e-cigarettes” at W1 and “e-products” (i.e., e-cigarettes, e-cigars, e-pipes, and e-hookah) at W2 and later waves. The current analyses treat e-cigarettes and ENDS the same. ^c^Other combustible includes traditional cigars, cigarillos, filtered cigars, pipe tobacco, and hookah tobacco; Smokeless tobacco includes smokeless tobacco, snus; Oral nicotine includes lozenges, discs, tablets, gum, toothpicks, dissolvable tobacco, and related products, but exclude nicotine replacement therapy. Nicotine pouches were asked about separately. Oral Nicotine/Nicotine Pouches were added at W7 only. ^d^ Continuous variable for adults only.
